# *TMEM106B* haplotypes show distinct associations with tau and TDP-43 pathologies in the aging brain

**DOI:** 10.64898/2026.08.11.26359568

**Authors:** Alex N. Salazar, Niccolò Tesi, Sven van der Lee, Frank Koopmans, Ka Wan Li, Susan K. Rohde, Maruelle C. Luimes, Annemieke J.M. Rozemuller, August B. Smit, Marc Hulsman, Henne Holstege

## Abstract

A central challenge in post-GWAS biology is determining how inherited variation within disease-associated loci shapes molecular mechanisms and clinical phenotypes. Here, we examined four previously identified *TMEM106B* haplotypes (T1-T4), defined by distinct combinations of coding, structural and regulatory variants. We integrated transcriptomic, proteomic, and neuropathological data from 1,299 individuals across two independent complementary aging cohorts. Although T2 and T3 both carry the p.Ser185 coding variant, they showed opposing associations with tau pathology, indicating that the surrounding haplotypic background modifies disease susceptibility. T3, which is enriched in cognitively healthy centenarians, was associated with lower tau pathology, lower C-terminal TMEM106B abundance, and reduced detection of an inflammatory microglial state, differing from the association pattern observed for T2. By contrast, T1 was associated with more extensive TDP-43 pathology, neuronal endolysosomal dysregulation, and increased C-terminal TMEM106B abundance. These findings identify haplotype-specific associations with differential proteinopathy burden, illustrating how haplotype-resolved analyses can connect GWAS signals to candidate molecular pathways.

## INTRODUCTION

With age, the human brain accumulates misfolded proteins linked to neurodegenerative disease, including Aβ and tau in Alzheimer’s disease (AD)^1,2^, α-synuclein in Parkinson’s disease^3^, and TDP-43 in limbic-predominant age-related TDP-43 encephalopathy (LATE)^4^. Yet, cognitively healthy centenarians reach at least 100 years of age with preserved cognition despite variable burdens of age-related neuropathology^5–7^. Genetic studies suggest that this resilience is partly mediated by enrichment for protective alleles and depletion of risk-increasing alleles, with strong effects at loci involved in endolysosomal and immune pathways^8^.

Recent proteomic and neuropathological analyses suggest that centenarian resilience is not defined solely by the absence of AD-related pathology. Instead, a subset of centenarians shows partial decoupling of Aβ and tau, with comparatively low tau pathology despite substantial amyloid burden^9^ (Rohde et al. 2026 submitted). This indicates that preserved cognition at extreme ages may depend on mechanisms that limit tau accumulation or spread downstream of amyloid. Identifying genetic factors that confer tau resilience is therefore central to understanding preserved cognition despite advanced age and neuropathological burden.

In this context, *TMEM106B* is of particular interest. One of the most prominently enriched genetic signals in cognitively healthy centenarians maps to the *TMEM106B* locus, where rs13237518-A is associated with reduced AD risk and more frequently carried by centenarians^8^ **(Table 1)**. This allele tags a common haplotype carrying serine at TMEM106B protein residue 185, whereas the reciprocal haplotype carries threonine **(Fig. 1a)**. The same linked alleles have also been associated with FTLD-TDP, LATE and hippocampal sclerosis, with the 185-Ser-tagged haplotype conferring reduced risk and the 185-Thr-tagged haplotype conferring increased risk^10–12^. Mechanistically, 185-Ser has been linked to reduced aggregation propensity of TMEM106B C-terminal fragments, whereas 185-Thr promotes fibril formation implicated in neurotoxicity and TDP-43 aggregation^13,14^. This supports a prevailing model in which 185-Ser is protective and 185-Thr is risk-increasing for TMEM106B- and TDP-43-related neurodegeneration.

**Fig. 1.**
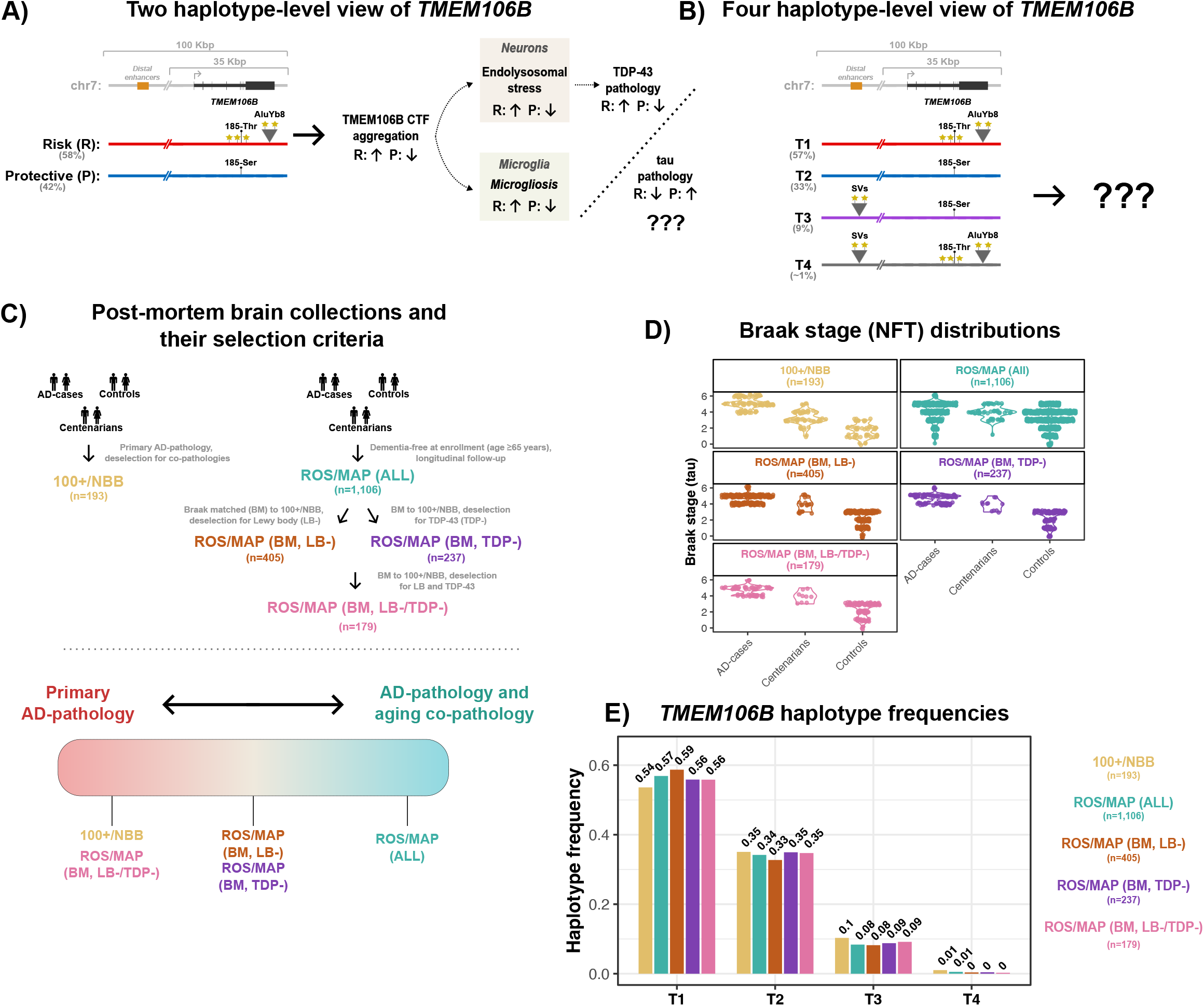
Haplotype-level framework and study design for resolving TMEM106B-associated proteinopathy associations. **(A)** Left: current two-haplotype view of the *TMEM106B* locus and its genetic features, contrasting the 185-Thr/AluYb8 and 185-Ser backgrounds. Right: the two-haplotype view explains associations with *TMEM106B* fibril formation, TDP-43 proteinopathy and cognitive decline, but remains contradictory for tau pathology^12–15^ **(Table 1)**. **(B)** Four-haplotype structure of the *TMEM106B* locus defined previously^16^, including T1, T2, T3 and the rare recombinant T4 haplotype. While this model resolves the genetic architecture beyond Thr185Ser, the functional consequences of these haplotypes remain unknown. **(C)** Overview of the two independent post-mortem brain collections analyzed in this study. Top: selection criteria for 100+/NBB and ROS/MAP. Bottom: schematic positioning of the collections along a spectrum from primary AD-related pathology to AD with frequent age-related co-pathologies. **(D)** NFT-Braak stage distributions across 100+/NBB and ROS/MAP. **(E)** *TMEM106B* haplotype frequencies across the analyzed collections.

**Table 1.**
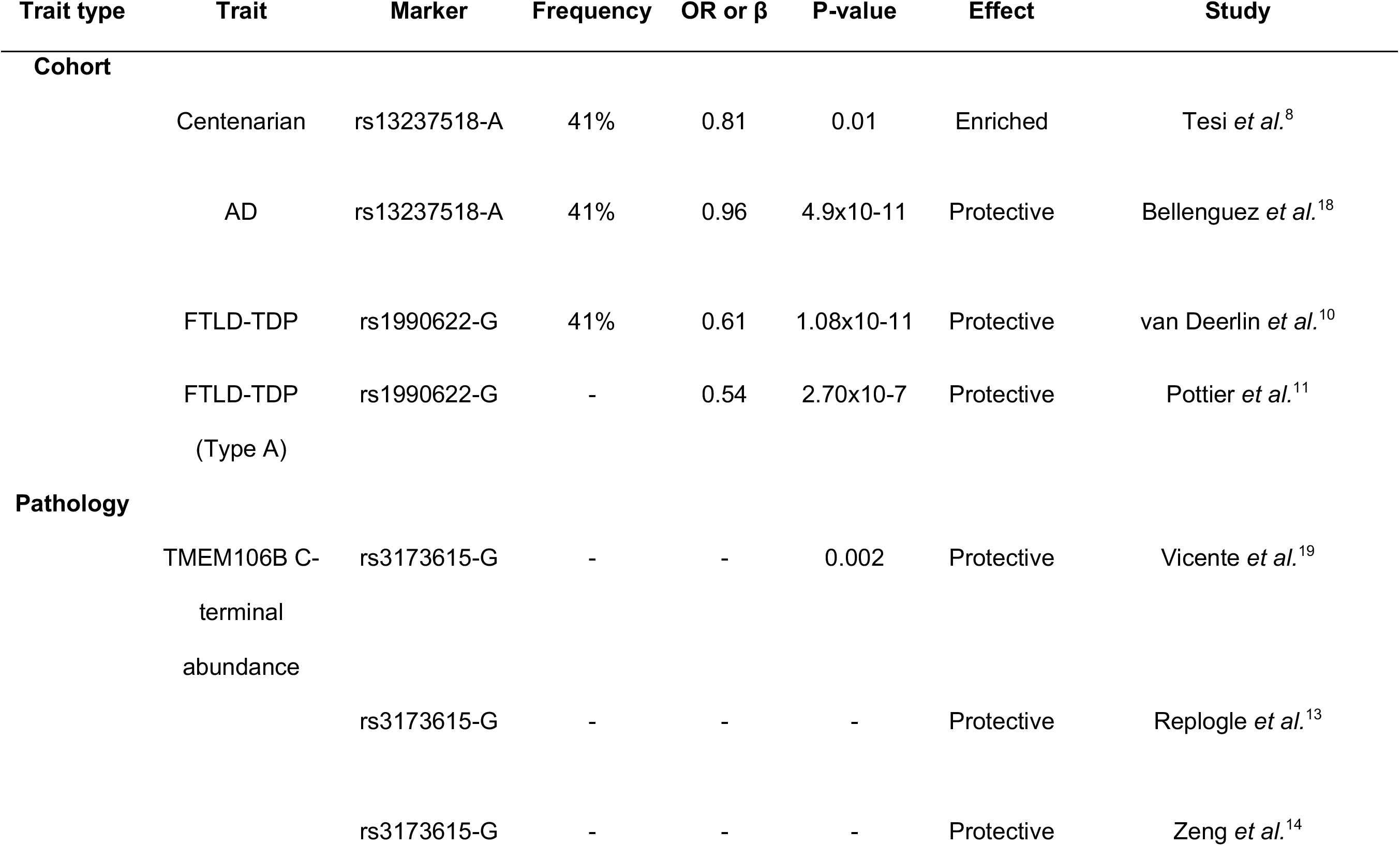
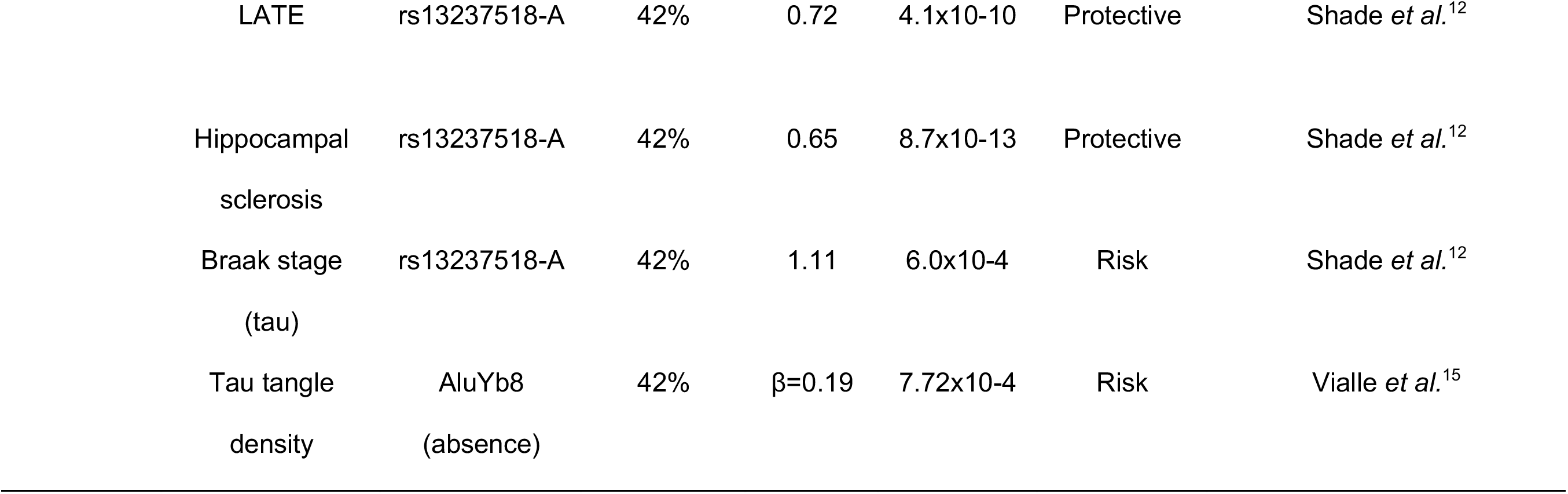
Prior associations of the *TMEM106B* haplotype across cohorts and pathologies.

However, this model does not explain all genetic associations at the locus **(Table 1)**. The same 185-Ser-tagged haplotype that appears protective for TDP-43-related disease has also been associated with greater tau burden and spread, whereas the reciprocal 185-Thr/AluYb8-linked haplotype has been associated with reduced tau pathology^12,15^ **(Table 1)**. If tau protection is a hallmark of preserved cognition at extreme age, then the single-variant model creates an apparent contradiction: centenarians are enriched for the 185-Ser allele linked to greater tau pathology, yet depleted for the reciprocal 185-Thr/AluYb8-linked allele associated with lower tau pathology.

We recently showed that, in addition to the haplotype characterized by Thr185Ser, the *TMEM106B* locus contains at least one further LD block that independently associates with AD at genome-wide significance^16^. Together, these variants define four haplotypes with distinct combinations of structural variants, coding SNPs and methylation patterns **(Fig. 1b)**. T1 carries 185-Thr in complete linkage with an AluYb8 insertion in the *TMEM106B* 3′UTR. T2 and T3 both encode 185-Ser, but T3 additionally contains multiple structural variants, including a ∼19 Kbp genomic rearrangement near distal regulatory elements. T4 is a relatively rarer recombinant haplotype carrying the 185-Thr/AluYb8 background in combination with the upstream structural variant cluster **(Fig. 1b)**.

These observations suggest that single-variant associations at *TMEM106B* may obscure distinct effects of the underlying haplotypes. Although T2 and T3 both encode 185-Ser, cognitively healthy centenarians are more strongly enriched for T3 (odds ratio for T3 [ORT3]=[1.28,1.42]) than for T2 (ORT2=[1.10,1.11]), suggesting that protection may not be explained by the coding substitution alone^16^. Conversely, T1 is depleted in centenarians (ORT1=[0.90,0.91]), despite prior associations with reduced tau spread, suggesting that associations with lower tau pathology do not necessarily indicate broader proteinopathy burden. Given *TMEM106B*’s role in endolysosomal homeostasis^13,14,17^, resolving haplotype-specific associations may clarify how this locus contributes to preserved cognition at extreme age.

Here, we integrated genetic, neuropathological, transcriptomic and proteomic data from 1,299 post-mortem brains spanning individuals with AD, non-demented controls and cognitively healthy centenarians aged 51-112 years. We evaluated whether T3 has a distinct association profile with tau and TDP-43 pathology relative to T2, despite their shared 185-Ser coding background; and further examined whether the other *TMEM106B* haplotypes showed concordant or contrasting associations across neuropathological and molecular outcomes. We show that these distinct haplotypes have divergent associations with tau and TDP-43 pathology, and identify cell-type-specific transcriptional and proteomic signatures implicating endolysosomal and microglial pathways. Together, these findings show how haplotype resolution can distinguish patterns of proteinopathy association that are obscured by single-variant models.

## RESULTS

### Dissection of haplotype-specific associations on neuropathological hallmarks in deeply phenotyped brain collections

To investigate haplotype-specific associations on neurodegenerative pathology, we analyzed two complementary post-mortem brain collections for which genotyping data was available: 100+/Netherlands Brain Bank (100+/NBB) collection^9^ and the Religious Orders Study and Memory and Aging Project (ROS/MAP) collection^20^ (**Fig. 1c**).

The 100+/NBB collection (n=193) comprises brain samples from neuropathologically confirmed AD-cases and age-matched (51-99) non-demented controls, collected through the Netherlands Brain Bank (NBB)^21^, as well as cognitively healthy centenarians (n=56) recruited through the 100-plus Study^5^. This collection was retrospectively assembled to maximize contrast in AD-related spreading of neurofibrillary tangles (NFTs) as measured by Braak staging^6^, while minimizing other neurodegenerative co-pathologies, including TDP-43 inclusions and α-synuclein pathology^9^. Accordingly, AD cases predominantly exhibit advanced Braak stages (≥IV), whereas non-demented controls show limited NFT pathology (Braak ≤III) **(Fig. 1c,d)**, contrasting early versus widespread limbic involvement of tau pathology. Centenarians were not preselected based on neuropathology and showed variable NFT spread, with Braak stages II-V^6,7^ **(Fig. 1c,d)**.

In contrast, ROS/MAP represents a prospective study of natural aging with longitudinal cognitive assessment and systematic neuropathological evaluation at autopsy^20^, capturing a broader spectrum of age-related pathologies^22^ **(Fig. 1C,D and Supplementary Fig. 1)**. Compared with 100+/NBB, ROS/MAP donors (n=1,106) show a broader continuum of tau pathology (p=5.81×10-3; FDR-adjusted q=0.008), and higher prevalence of TDP-43 and Lewy body pathology (TDP-43: p=7.30×10-9; q=7.30×10-8; Lewy body: p=3.69×10-6; q=1.23×10-5) **(Supplementary Fig. 1)**. The 100+/NBB collection also contained more female donors than ROS/MAP (χ2-test: p=0.01; q=0.013) **(Supplementary Fig. 1)**, particularly among AD cases (100+/NBB: 85.1%, n=74; ROS/MAP: 68.0%, n=302).

We assigned *TMEM106B* haplotype dosage values (0, 1 or 2) corresponding to non-carriers, heterozygous carriers, and homozygous carriers, respectively, using previously defined SNP marker combinations^16^. Haplotype frequencies were comparable between 100+/NBB and ROS/MAP **(Fig. 1e)**, and consistent with the general Dutch population: T1 (53.6% and 56.9%, respectively), T2 (35.1%, 34.2%), T3 (10.3%, 8.4%) and T4 (1.0%, 0.5%).

Overall, these differences in pathology composition motivated *in silico* subsetting of ROS/MAP to better match the 100+/NBB pathology profile **(Fig. 1c-e)**, enabling direct comparison of haplotype associations across the collections.

### The T3 haplotype in TMEM106B is associated with lower tau pathology in 100+/NBB

We first assessed the association between haplotype dosage and overall tau spreading (NFT-Braak stage) in the whole 100+/NBB collection, using a joint model including all haplotypes with T2 (185-Ser only) as reference, and adjusted for age at death, sex, and global amyloid burden. T4 was included in the model but excluded from downstream interpretation because of its low frequency. We found that T3 haplotype (185-Ser+SVs) was associated with lower NFT Braak stage (OR=0.34; p=0.003; FDR-adjusted q=0.015) **(Supplementary Fig. 2a)**.

Braak staging reflects the regional spread of neurofibrillary tangles from entorhinal and hippocampal regions (stages 0-II), through the limbic system (III-IV), to neocortical involvement (V-VI) **(Fig. 2a)**. Because 100+/NBB spans these stages **(Fig. 1d)**, we next allowed haplotype associations to differ at each stage transition. Relative to T2, T3 was associated with greater odds of remaining at Braak 0-II rather than progressing to stages III-VI (OR=5.94 [1.87,18.89]; p=0.003, q=0.026), but not with the distinction between stages 0-IV and neocortical stages V-VI (OR=1.56 [0.40,4.45], p=0.402, q=0.502) **(Fig. 2b,c)**. Because T3 was relatively less common than T1 and T2, we confirmed the first association using bias-reduced logistic regression, which similarly showed lower odds of Braak III-VI among individuals carrying T3 (OR=0.29 [0.10,0.81], p=0.018) **(Supplementary Fig. 2b)**. These findings suggest that T3 is preferentially associated with brains lacking limbic tau pathology, rather than with reduced progression from limbic to neocortical stages.

**Fig. 2.**
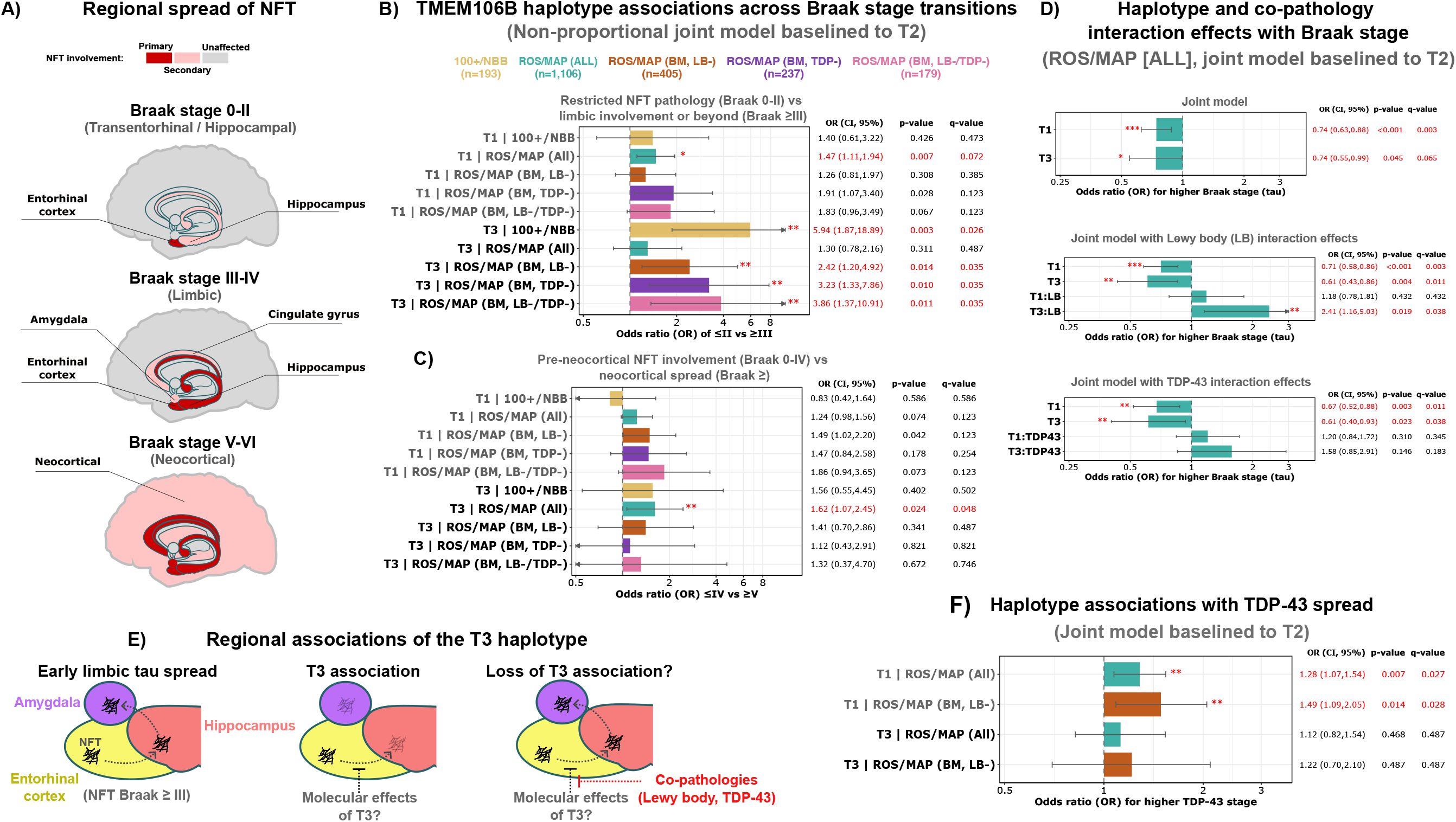
*TMEM106B* haplotype associations with tau and TDP-43 spread. **(A)** Neurofibrillary tangle (NFT) spread across the regional anatomy of the limbic system and neocortex based on Braak staging. **(B-D)** Joint association models testing *TMEM106B* haplotype carriership on tau or TDP-43 spread across all post-mortem brain collections and subsets, each with a unique color as defined in Fig. 1C. Colored bars indicate estimated odds ratio (OR), and black horizontal lines represent 95% confidence intervals. Asterisks indicate FDR-adjusted q-values (10% threshold): *** q < 0.01; ** q < 0.05; * q < 0.1. Red text denotes an association with statistical significance (q < 0.1). **(B)** Association of haplotype carriership with restricted NFT pathology (Braak ≤II) vs limbic involvement (Braak ≥III). **(C)** Haplotype associations with NFT pre-neocortical spread (Braak ≤IV) vs neocortical spread (Braak ≥V). **(D)** Interaction effects between haplotypes and co-pathologies in the full ROS/MAP collection. **(E)** Possible interpretation of T3’s association with lower limbic tau involvement. **(F)** Association of haplotype carriership with overall TDP-43 spread in ROS/MAP.

We recently observed that the spatiotemporal spreading of aggregated tau tangles across brain regions, as reflected by Braak staging, is an independent aspect of tau-progression from the increased molecular *abundance* of tau^9^. Therefore, we leveraged the 100+/NBB collection for its availability of previously generated cell-type corrected bulk LC-MS/MS proteomics data from the middle temporal gyrus^9^ **(Fig. 3a)**, which is affected by phosphorylated tau spreading at Braak stages III-IV.

**Fig. 3.**
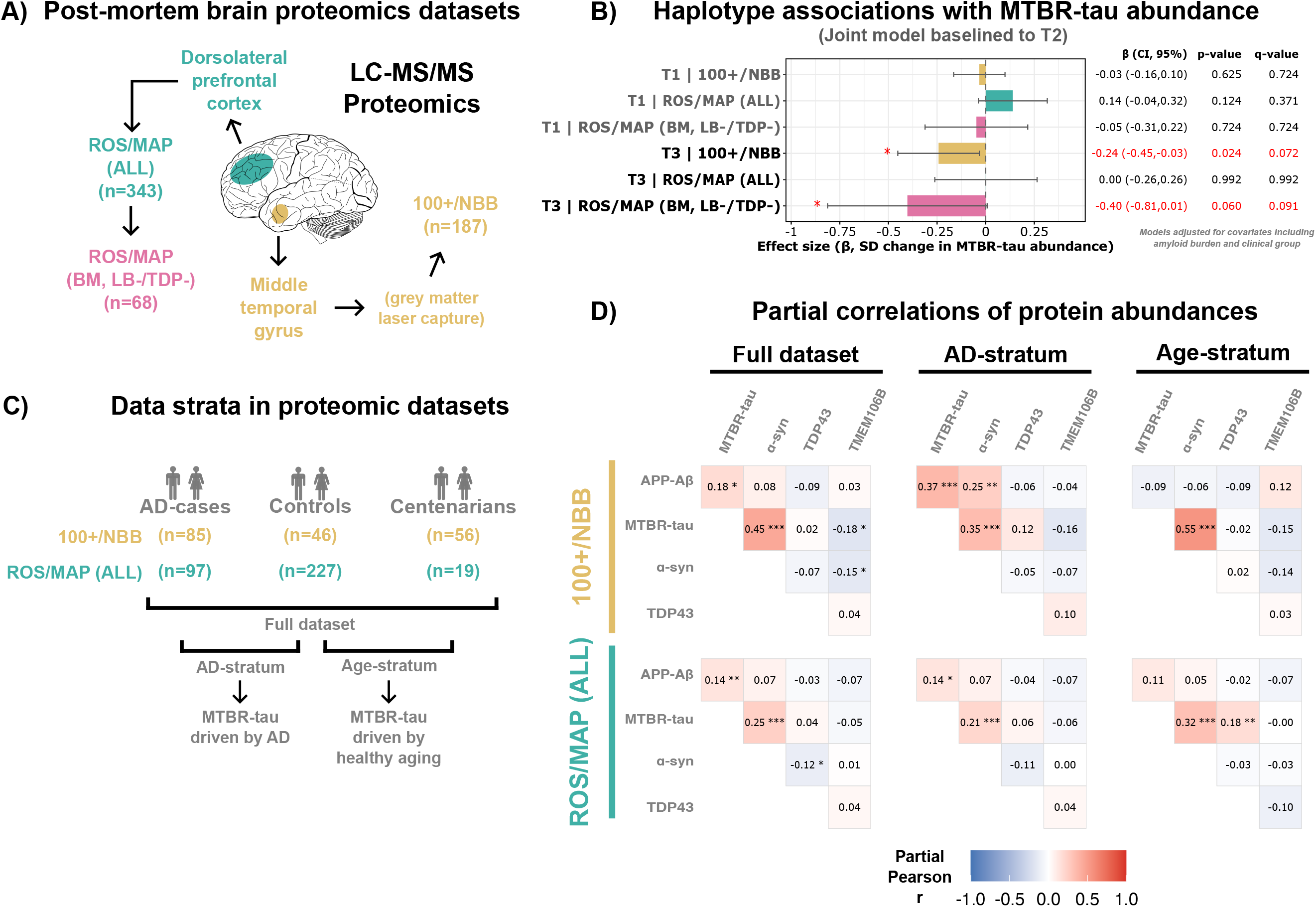
*TMEM106B* haplotype associations with proteomic tau abundance, along with correlations of other proteins. **(A)** LC-MS/MS proteomic datasets were previously generated in brain donors of 100+/NBB (n=187, middle temporal gyrus^9^) and ROS/MAP (n=343, dorsolateral prefrontal cortex^23^), along with in silico subsets of ROS/MAP with co-pathologies matched to 100+/NBB (n=68; Braak stage matched [BM], exclusion of brains with Lewy body [LB-] and TDP-43 [TDP43-]). Cell-type fraction correction of proteomic abundances was only possible in 100+/NBB^9,23^. **(B)** Association of *TMEM106B* haplotypes with protein abundance of the microtubule binding region of tau (MTBR-tau) across data collections and subsets; adjusted for covariates including amyloid burden and clinical group (see Methods). To ensure comparability across models, MTBR-tau abundance was z-score standardized. Colored bars indicate estimated effects as changes in SD units of MTBR-tau abundance, and black horizontal lines represent 95% confidence intervals. **(C)** Both 100+/NBB and ROS/MAP can be partitioned to disentangle AD-related vs age-related effects by defining AD-stratum (AD-cases vs controls) and Age-stratum (controls vs centenarians)**. (D)** Partial correlations in 100+/NBB and ROS/MAP proteomic datasets across each strata. **(B, D)** Asterisks indicate FDR-adjusted q-values (10% threshold): *** q < 0.01; ** q < 0.05; * q < 0.1. Red denotes statistical significance (q < 0.1).

Relative to T2 and after adjustment for covariates including age at death, sex, local amyloid burden, and clinical group (AD case, control or centenarian), T3 was associated with lower abundance of soluble peptides from the aggregation-prone microtubule-binding region of tau (MTBR-tau) (β=−0.24; p=0.024; q=0.072) **(Fig. 3b)**.

Together, these findings highlight that T3 showed the strongest association with lower tau pathology in 100+/NBB, across both tau spread and MTBR-tau abundance.

### The association between T3 and lower tau pathology is also observed in ROS/MAP but varies by co-pathology context

We observed a significant, but attenuated association between T3 and lower overall tau spread in the full ROS/MAP collection (NFT Braak stage: OR=0.74, 95% CI=[0.55,0.99], p=0.045, q=0.057) **(Supplementary Fig. 2a)**. Because 100+/NBB was retrospectively assembled to maximize contrast in AD-related tau pathology while depleting Lewy body and TDP-43 co-pathologies, we generated *in silico* ROS/MAP subsets that progressively aligned with the 100+/NBB selection strategy: a Braak-matched subset (BM) followed by exclusion of brains with Lewy body pathology (BM, LB−; n = 405), TDP-43 pathology (BM, TDP43−; n = 237), and both co-pathologies (BM, LB−/TDP43−; n = 179) **(Fig. 1c)**.

The T3 association strengthened across these subsets and was largest after excluding both co-pathologies (BM, LB-/TDP43-: OR=0.45 [0.22,0.94], p=0.034, q=0.057) **(Supplementary Fig. 2a)**. The subsets showed the same stage-specific pattern observed in 100+/NBB, with the strongest association again in the BM, LB-/TDP43-subset: T3 was associated with greater odds of Braak 0-II rather than III-VI (OR=3.86 [1.37,10.91], p=0.011, q=0.035), but not with Braak 0-IV rather than V-VI (OR=1.32 [0.37,4.70], p=0.672, q=0.746) **(Fig. 2b,c)**. Bias-reduced logistic regression similarly showed lower odds of Braak III-VI among T3 carriers across all subsets, with the strongest association in BM, LB-/TDP43-(OR=0.34 [0.13,0.85], p=0.021) **(Supplementary Fig. 2b)**. Thus, after minimizing co-pathology burden, ROS/MAP also showed an association between T3 and lower odds of limbic tau involvement and beyond.

Interaction models in the full ROS/MAP collection showed that co-pathologies modified the T3-tau association **(Fig. 2d)**. Relative to T2, T3 was associated with lower Braak stage in brains without Lewy body pathology or TDP-43 pathology (T3LB-: OR=0.61; p=0.004; q=0.011; T3TDP43-: OR=0.61; p=0.023; q=0.038). This association was significantly attenuated in the presence of Lewy body pathology (T3xLB: OR=2.41; p=0.019; q=0.038), whereas the interaction with TDP-43 pathology was not significant (T3xTDP43: OR=1.58; p=0.146; q=0.183). Lewy body interaction remained significant after additional adjustment for study cohort and clinical group (T3LB-: OR=0.63; p=0.008; T3xLB: OR=2.23; p=0.033; T3TDP43-: OR=0.66; p=0.033; T3xTDP43: OR=1.51, p=0.192). Thus, the association between T3 and lower tau spread was strongest in brains without Lewy body or TDP-43 co-pathology.

Next, we tested whether this association was also detectable at the molecular level. In ROS/MAP LC-MS/MS proteomic data from the dorsolateral prefrontal cortex (n=343)^23^ **(Fig. 3a)**, T3 carriers showed lower MTBR-tau abundance in the Braak-matched LB-/TDP43-subset after covariate adjustment, including study cohort and clinical group (n=68; β=−0.40; p=0.061; q=0.091) **(Fig. 3b)**.

Finally, we contextualized the proteomic associations on MTBR-tau abundance by defining three separate data strata in both 100+/NBB and ROS/MAP proteomic datasets: full dataset, AD-stratum (AD-cases vs controls), and Age-stratum (controls vs centenarians) **(Fig. 3c)**. Partial correlations of Aβ, MTBR-tau, TDP-43, TMEM106B, and α-synuclein abundances across these strata highlighted that MTBR-tau abundance was significantly correlated with α-synuclein abundance in both datasets (100+/NBB: r=0.45, p=9.72×10-11, q=9.72×10-10; ROS/MAP: r=0.25, p=4.00×10-6, q=4.00×10-5), which was strongest in the Age-stratum (100+/NBB: r=0.55, p=1.93×10-9, q=1.93×10-8; ROS/MAP: r=0.32, p=2.52×10-7, q=2.52×10-6) **(Fig. 3d)**.

Together, these findings show that the association between T3 and lower tau pathology was also observed in ROS/MAP, was strongest in co-pathology-depleted subsets, and differed significantly by Lewy body (α-synuclein) status.

### The T1 haplotype shows a less consistent association with lower tau pathology than T3

We next asked whether the T1 haplotype, which carries 185-Thr and the AluYb8 insertion, showed an association with tau spread comparable to T3. In the full ROS/MAP cohort, T1 was associated with lower overall NFT Braak stage (OR=0.74; p=5.82×10-4; q=0.003) **(Supplementary Fig. 2a)**, with an effect size similar to T3. However, subsequent analyses indicated that the T1 association was both less consistent and distinct from the T3 pattern.

First, T1 was not associated with overall tau pathology in 100+/NBB **(Supplementary Fig. 2a)**. Second, after Braak matching and co-pathology reduction in ROS/MAP, T1 was associated with lower Braak stage but showed consistently weaker effect estimates than T3 (T1: OR=0.58-0.74; T3: OR=0.45-0.58) **(Supplementary Fig. 2a)**. Third, unlike T3, T1 did not show a consistent association at the transition from Braak stages 0-II to III-VI across these subsets **(Fig. 1b,c)**. Across the Braak-matched subsets, T3 showed consistently stronger associations with remaining at Braak stages 0-II (OR=2.42-3.86) than T1 (OR=1.26-1.91), and the T1 association remained significant after FDR correction only in the full ROS/MAP collection. Fourth, the T1 association was not significantly modified by Lewy body or TDP-43 co-pathology **(Fig. 1d)**. Finally, T1 showed no significant association with MTBR-tau abundance in 100+/NBB, the full ROS/MAP proteomic dataset, or the Braak-matched LB-/TDP43-ROS/MAP proteomic subset **(Fig. 3b)**.

### T1 is associated with greater TDP-43 spread

Given *TMEM106B*’s prior implication in TDP-43 proteinopathies **(Table 1)**, we next examined whether the haplotypes were associated with TDP-43 pathology. Because the 100+/NBB collection was assembled to minimize TDP-43 co-pathology, we focused on the semi-quantitative TDP-43 staging available in ROS/MAP (n=1,019).

Consistent with prior findings **(Table 1)**, T1 (185Thr/AluYb8) was associated with increased TDP-43 spread relative to T2 carriers (OR=1.28; p=0.007; q=0.027) **(Fig. 2f)**. This effect was stronger in the Braak-matched ROS/MAP subset depleted of Lewy body co-pathology (BM, LB-) (OR=1.49, p=0.014, q=0.028) **(Fig. 2f)**.

However, unlike T3, Lewy body co-pathology did not significantly modulate the association between T1 and increased TDP-43 spread in the full ROS/MAP collection **(Supplementary Fig. 2c)**. In the proteomic data, T1 was not significantly associated with TDP-43 abundance in 100+/NBB nor ROS/MAP **(Supplementary Fig. 2d)**. We also observed no significant effects of the T3 haplotype on TDP-43 spread or abundance **(Fig. 2F and Supplementary Fig. 2c,d)**.

As each individual carries two *TMEM106B* haplotypes, we next tested whether specific haplotype combinations, i.e. genotypes, associate with tau (NFT-Braak stage) and TDP-43 (TDP-43 stages) spread using a joint model with sum-to-zero encoding. This allowed each genotype to be interpreted as a deviation from the mean effect across all genotype combinations **(Supplementary Fig. 3)**. T3/T3 carriers had the lowest estimated tau and TDP-43 stages, although this finding was limited by the small number of homozygous T3 carriers (n=14) **(Supplementary Fig. 3)**.

### TMEM106B haplotypes are associated with downstream transcriptional and proteomic variation in a local four-gene promoter hub

Integration of publicly available proximity ligation-assisted ChIP-seq (PLAC-seq) data^24^ suggested that the *TMEM106B* locus forms a four-gene promoter hub **(Fig. 4a,b)**, with potential coordinated regulation of *TMEM106B*, *THSD7A*, *VWDE*, and *C7orf78* by *TMEM106B* haplotypes.

**Fig. 4.**
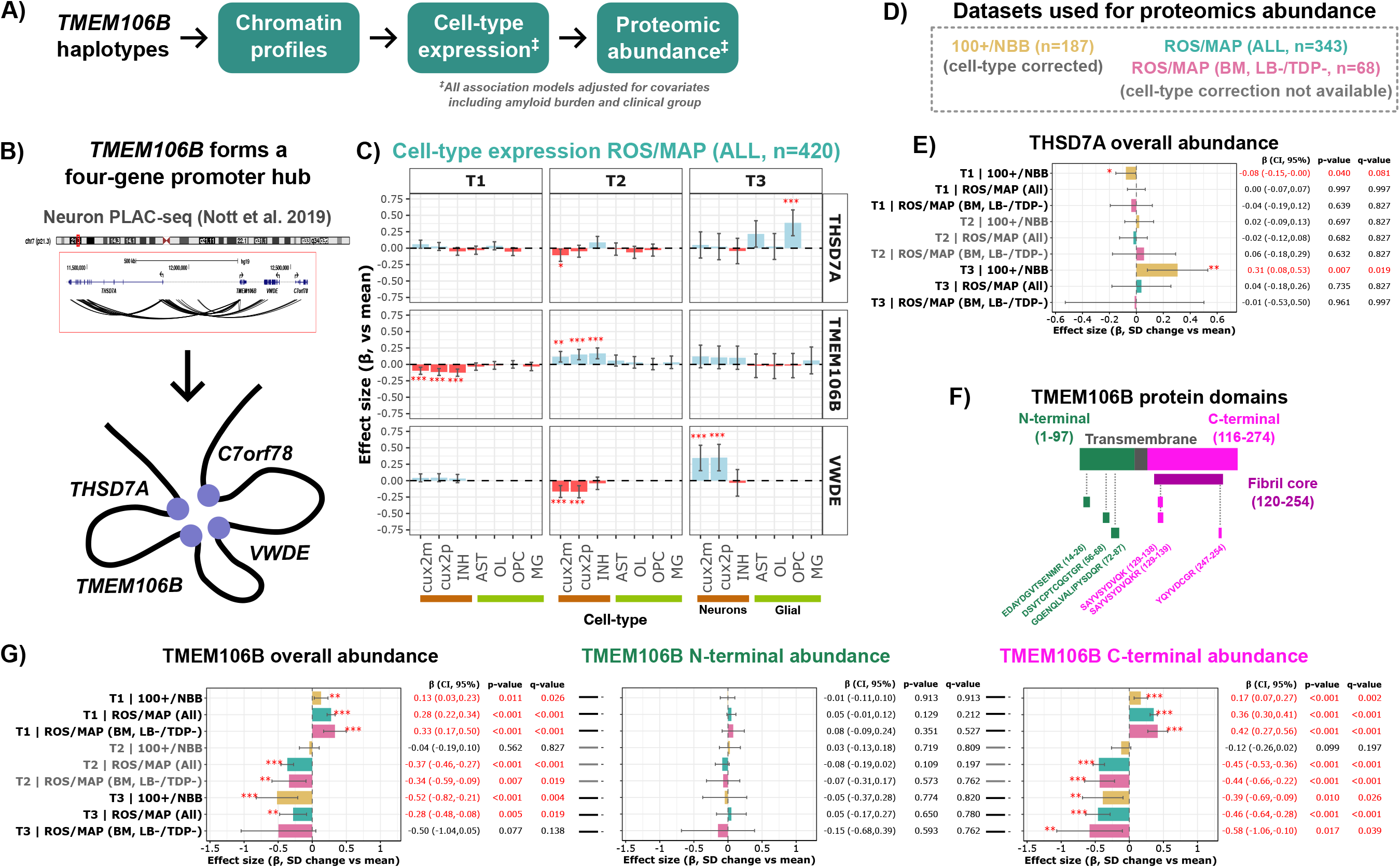
Multi-modal integration enables *TMEM106B* haplotype associations across single-nucleus transcriptomics and bulk proteomics. (A) Overview of the multi-modal integration framework used to evaluate cis-regulatory associations of *TMEM106B* haplotypes. **(B)** H3K4me3-anchored PLAC-seq data from Nott *et al.*^24^ suggest that, at least in neurons, *TMEM106B* participates in a four-gene promoter-centered interaction hub. **(C-G)** Regression models associating *TMEM106B* haplotypes in single-nucleus expression and bulk proteomic datasets; adjusted for covariates including amyloid burden and clinical group. Asterisks and text indicate Meff-adjusted^25^ q-values (10% threshold): *** q < 0.01; ** q < 0.05; * q < 0.1. Red denotes statistical significance (q < 0.1). **(C)** Haplotype associations based on single-nucleus RNA-sequencing data from ROS/MAP (n=420, dorsolateral prefrontal cortex). Colored bars indicate estimated effects as changes in rank-normalized gene expression (blue: increased expression [i.e. beta > 0]; red: decreased expression), with black horizontal lines representing 95% confidence intervals. X-axis depicts cell-types by neurons (excitatory [CUX2- and CUX2+], and inhibitory [INH]) and glial (astrocytes [AST], oligodendrocytes [OL], oligodendrocytes precursor cells [OPC], microglia [MG]). **(D)** LC-MS/MS proteomic datasets as introduced in Fig. 3. The 100+/NBB dataset was cell-type fraction corrected^9^, whereas correction was not available for ROS/MAP^23^. **(E)** Haplotype associations with overall THSD7A protein abundance. **(F)** Overview of TMEM106B protein domains; numbers indicate amino acid residues defining domain boundaries based on Jiang *et al.*^26^. The peptides shown were used to estimate overall and domain-specific abundance. **(G)** Haplotype associations with overall (left), N-terminal (middle), and C-terminal (right) TMEM106B protein abundance.

Using single-nucleus RNA-seq data from 420 dorsolateral prefrontal cortices from ROS/MAP^27^, we found that *TMEM106B* haplotypes differentially associate with expression of *TMEM106B*, *THSD7A*, and *VWDE* across neuronal and glial cell types **(Fig. 4c)**. To estimate mutually adjusted haplotype associations, we fitted a joint model with sum-to-zero encoding, enabling direct comparisons across haplotypes **(see Methods)**. T1 was associated with lower expression of *TMEM106B* in inhibitory and excitatory neurons (β=[-0.13,-0.10], p≤2.02×10-4, q≤8.96×10-4). Conversely, T2 was associated with upregulation of *TMEM106B* in the same cell-types (β=[0.14,0.19], p≤8.10×10-4, q≤0.004) **(Fig. 4c)**. T3 did not significantly alter *TMEM106B* expression **(Fig. 4c)**, and T4 was excluded from interpretation due to its low frequency.

These transcriptional associations were variably reflected at the protein level using bulk proteomics from the 100+/NBB (temporal cortex^9^) and ROS/MAP (frontal cortex^23^) proteomics collections **(Fig. 4d-g)**, with cell-type fraction correction available only in 100+/NBB^9,23^. Although T2 was associated with higher *TMEM106B* RNA expression, T2 associated with *lower* overall TMEM106B protein abundance in ROS/MAP (ALL: β=-0.37, p=9.50×10-15, q=8.55×10-14; BM, LB-/TDP43-: β=-0.34, p=0.007, q=0.019), which was driven by the C-terminal fragment, with no effect on N-terminal abundance. However, T2 did not associate with TMEM106B abundance in 100+/NBB (β=-0.04 p=0.562, q=0.827) **(Fig. 4g)**. Conversely, while T1 associated with lower *TMEM106B* RNA expression, it associated with higher TMEM106B protein abundance in both ROS/MAP (ALL: β=0.28, p=8.36×10-20, q=1.50×10-18; BM, LB-/TDP43-: β=0.33, p=8.41×10-5, q=5.05×10-4) and 100+/NBB (β=0.13, p=0.011, q=0.026), again primarily driven by the C-terminal portion **(Fig. 4g)**.

Despite no significant effects on *TMEM106B* transcription, T3 was associated with lower TMEM106B protein abundance in both ROS/MAP (ALL: β=-0.28; p=0.005; q=0.019; LB-/TDP43-: β=-0.50; p=0.077; q=0.138) and 100+/NBB (β=-0.52; p=8.62×10-4; q=0.004), also primarily driven by the C-terminal portion **(Fig. 4g)**.

We additionally observed coordinated transcriptional and proteomic effects on *THSD7A* and *VWDE* **(Fig. 4c)**. T3 was associated with higher *THSD7A* expression in OPCs (β=0.39, p=8.49×10-5, q=3.77×10-4) and with higher *VWDE* expression in excitatory neurons (Cux2−: β=0.35, p=3.13×10-4, q=0.001; Cux2+: β=0.36, p=4.18×10-4, q=0.002). T3 was associated with higher THSD7A protein abundance in 100+/NBB (β=0.31, p=0.007, q=0.019), but not in ROS/MAP (ALL: β=0.04, p=0.735, q=0.827; LB-/TDP43-: β=-0.01, p=0.961, q=0.997). T2 showed opposing transcriptional effects on *THSD7A* and *VWDE*, without detectable effects at the protein level **(Fig. 4c-e)**. VWDE peptides, and *C7orf78* transcripts and peptides were not detected.

Finally, in sensitivity analyses additionally adjusting for local TMEM106B, THSD7A, and α-synuclein abundance, T3 remained nominally associated with lower MTBR-tau in 100+/NBB (β=−0.21, p=0.037); while the Braak-matched, LB-/TDP43-ROS/MAP subset showed a similarly sized effect (β=−0.34, p=0.108), although it was not nominally significant. Thus, further adjustment for these measured protein abundances did not eliminate the direction or magnitude of the T3’s association.

### TMEM106B haplotypes are associated with cell-type-specific expression patterns and microglial-state prevalence

Given *TMEM106B*’s role in endolysosomal homeostasis^13,14,17^, we tested whether its haplotypes associate with cell-type-specific expression of 1,383 endolysosomal and vesicle-trafficking genes using single-nucleus data from ROS/MAP (n=420) **(Fig. 5a and Supplementary Table 1)**. Using marginal regression models adjusted for age, sex, post-mortem interval, ancestry, global amyloid burden, study cohort and clinical group (e.g. AD-case, control, or centenarian), T1 showed significant neuronal associations with 445 genes (FDR-adjusted q<0.01), with 57% (n=254) showing decreased expression **(Supplementary Table 2)**. In contrast, T2 was significantly associated with neuronal expression of 254 genes, 66% (n=167) showing increased expression **(Fig. 5a)**. T1 and T2 showed broadly opposing effects on expression direction, particularly in CUX2+ excitatory neurons. Among their 30 strongest associations, 20 cell type-gene associations overlapped, all with opposing effect directions **(Supplementary Table 2)**, consistent with their reciprocal haplotypic backgrounds. T3 was associated with six genes distributed across neuronal and non-neuronal cell types, whereas T4 was excluded due to low frequency.

**Fig. 5.**
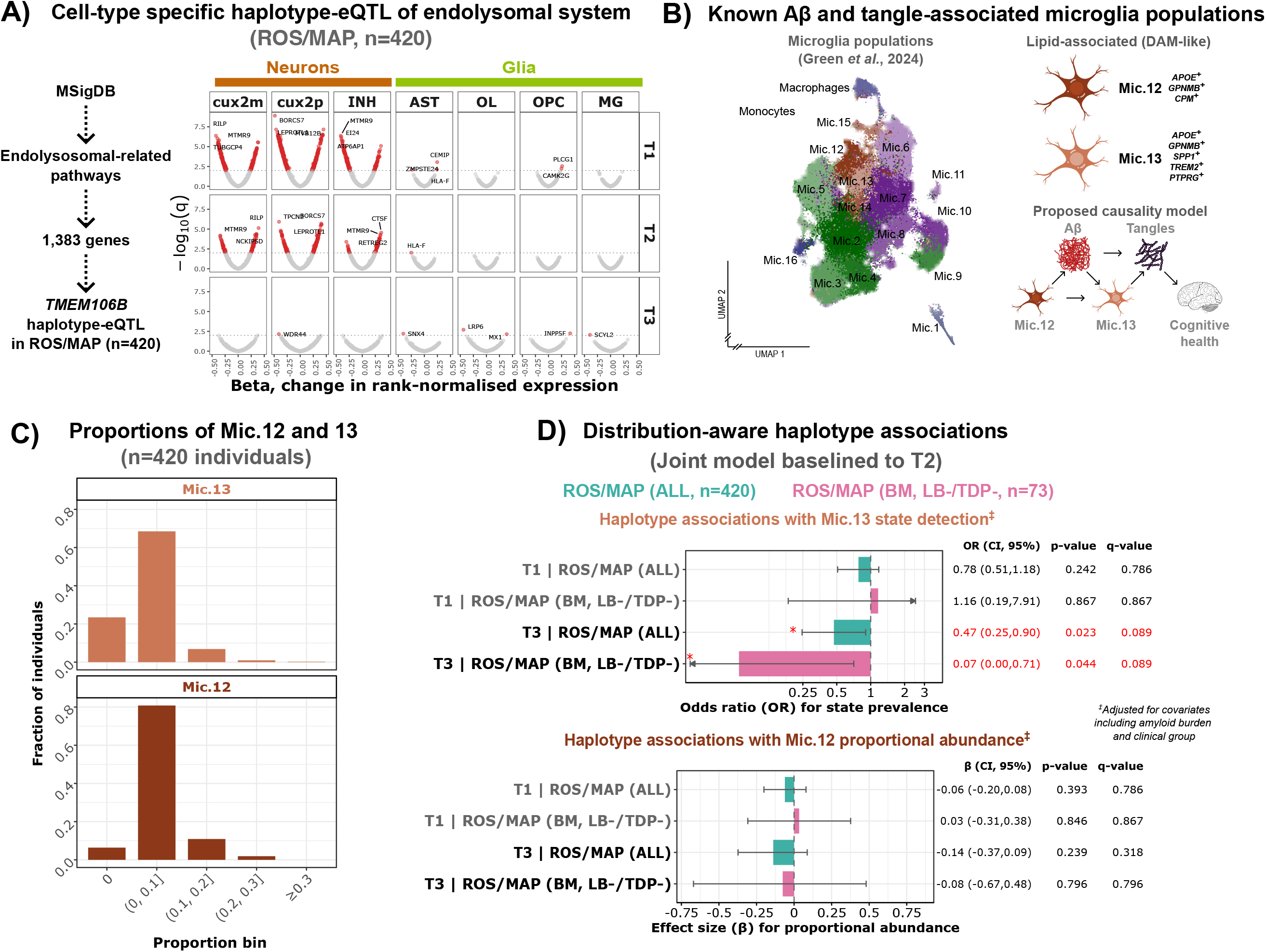
*TMEM106B* haplotype associations with endolysosomal gene expression and microglial states. **(A)** Volcano plots showing haplotype-specific effects on cell-type-specific gene expression using a curated set of 1,383 genes involved in endolysosomal pathways (MsigDB; **Supplementary Table 1**), in available single-nucleus data from ROS/MAP (n=420). X-axis: effect size; Y-axis: FDR-adjusted q-value. Red points indicate significantly differentially expressed genes (FDR-adjusted q-value < 0.01). Annotated genes represent the top ten differentially expressed genes per cell type and haplotype. **(B)** Left: UMAP projection of annotated microglial subpopulations from ROS/MAP single-nucleus data (adapted from Green *et al.*^27^). Right: Mic.12 and Mic.13 microglia states implicated in Alzheimer’s disease progression along with proposed causality model^27^. **(C)** Distribution of the proportions of Mic.12 and Mic.13 populations among individuals with available microglia state proportion estimations in ROS/MAP (n=420). **(D)** Associations of *TMEM106B* haplotypes on detection or proportional abundance of the Mic.12 and Mic.13 microglia populations **(Methods and Supplementary Results).** Asterisks and text indicate FDR-adjusted q-values: *** q < 0.01; ** q < 0.05; * q < 0.1. Red denotes statistical significance (q < 0.1).

*BORCS7* was the strongest T1- and T2-associated hit and showed reciprocal effects between haplotypes in CUX2+ neurons (T1: β=−0.49, p=5.40×10−14, q=1.24×10−9; T2: β=0.40, p=3.89×10−9, q=2.01×10−6). *BORCS7* is a component of the BORC complex, which promotes kinesin-dependent lysosomal transport into neuronal processes^28^. Reduced *BORCS7* expression with T1 may therefore limit the distal trafficking and release of tau-containing endolysosomal vesicles, whereas increased expression with T2 may facilitate their transport and propagation. Consistent with this mechanistic possibility, T1 was associated with lower CD81 expression in CUX2− (β=−0.29, p=3.04×10−5, q=0.001), CUX2+ (β=−0.40, p=5.86×10−9, q=2.60×10−6) and inhibitory neurons (β=−0.36, p=5.49×10−8, q=1.22×10−5), whereas T2 was associated with higher CD81 expression in CUX2+ (β=0.30, p=3.45×10−5, q=0.001) and inhibitory neurons (β=0.29, p=4.18×10−5, q=0.001). CD81 regulates extracellular-vesicle biogenesis and has been implicated in the packaging of tau fibrils^29^. These reciprocal effects suggest that reduced endolysosomal trafficking and vesicular tau packaging may contribute to the association between T1 and lower tau spread, whereas increased activity of these pathways may contribute to the opposing association observed with T2.

Because microglial responses have been implicated in tau propagation^27,30^, we next tested whether *TMEM106B* haplotypes associate with microglial state composition available in the single-nucleus data from ROS/MAP (n=420)^27^. We focused on Mic.12 and Mic.13, two AD-associated microglia populations previously linked with AD progression^27^ **(Fig. 5b)**. Mic.12 is an amyloid-responsive state marked by *CPM* and enriched for complement and phagocytic activation markers, including *APOE* and *GPNMB*; whereas Mic.13 is marked by *PTPRG*, *TREM2* and *SPP1*, and is positioned downstream of amyloid and more closely linked to tau burden and cognitive decline **(Fig. 5b)**.

Using a joint regression model relative to T2 and adjusting for the same covariate set as in the above haplotype-eQTL analysis, we found that T3 carriership was associated with reduced prevalence of Mic.13 state in the full ROS/MAP single-nucleus dataset (n=420) (OR=0.47, p=0.023, q=0.089), which was further amplified in the Braak matched, Lewy body and TDP-43 depleted subset (n=73) (OR=0.07, p=0.044, q=0.089) **(Fig. 5c,d)**. Sensitivity analyses using alternative population proportion-modeling approaches yielded concordant associations **(Supplementary Results)**. No significant association was observed for the other haplotypes or with Mic.12. These findings suggest that the association of T3 with lower tau pathology may involve reduced engagement of a specific disease-associated microglia state, rather than a broad suppression of amyloid-responsive microglial activation.

Finally, while T3 and T1 significantly associated with reduced local tau tangle density in the midfrontal cortex among all individuals in the single-nucleus data from ROS/MAP (T1: β=-0.15, 95% CI=[-0.30,-0.01], p=0.034; T3: β=-0.33, 95% CI=[-0.56,-0.10], p=0.006), only T3 remained significant after adjustment for Mic.13 state activation (T1: β=-0.10, 95% CI=[-0.25,0.04], p=0.151; T3: β=-0.27, 95% CI=[-0.50,-0.04], p=0.023), indicating that Mic.13-related changes do not fully explain the T3’s association with lower tau tangle density.

Overall, these findings suggest that *TMEM106B* haplotypes may be associated with tau through, or alongside, differences in neuronal and microglial states.

## DISCUSSION

Our study shows that the associations of *TMEM106B* locus with neurodegenerative traits are best understood at the haplotype level. By integrating genetic, transcriptomic, proteomic and neuropathological data across two independent post-mortem brain collections, we find that four *TMEM106B* haplotypes (T1-T4) differentially associate with tau, TDP-43 and TMEM106B proteinopathies. The associations suggest two partially opposing patterns: one related to C-terminal TMEM106B accumulation and TDP-43 vulnerability, and another marked by reduced tau burden. The key novel insight is the identification of T3 as the haplotype underlying this latter association. Although T2 (185-Ser) and T3 (185-Ser+SVs) both encode 185-Ser, T3 was associated with reduced tau spread and lower MTBR-tau abundance, whereas T2 showed the opposite pattern. The T3 association was strongest at the threshold for limbic involvement, raising the possibility that T3 marks a genetic background less permissive to the extension of tau pathology into limbic regions.

This haplotype-level view helps resolve previously paradoxical associations at the *TMEM106B* locus **(Fig. 1a and Table 1)**. At the variant level, the 185-Thr allele and presence of the AluYb8 insertion have been associated with increased risk for TDP-43/TMEM106B proteinopathy and cognitive decline, but also with protection against tau pathology^12,15^. Indeed, T1 carries the 185-Thr and AluYb8 alleles and was associated here with increased TDP-43 spread and higher C-terminal TMEM106B protein abundance, yet also showed a tau-protective association in ROS/MAP. As summarized in Fig. 6, we speculate that these opposing effects reflect two parallel processes: *TMEM106B* fibril accumulation may slow endolysosomal vesicle trafficking and thereby reduce tau propagation, while also increasing vulnerability to the accumulation of TDP-43 pathology; in parallel, increased TDP-43 fibril accumulation may further limit tau seeding or propagation via direct interactions^31^ and/or slowing of endolysosomal trafficking. Since T1 is depleted in cognitively healthy centenarians^16^, this route of tau reduction is unlikely to translate into a broadly favorable aging profile. T1 can therefore be interpreted as a trade-off haplotype, in which reduced tau propagation may occur as a consequence of increased TMEM106B and TDP-43 proteinopathy.

**Fig. 6.**
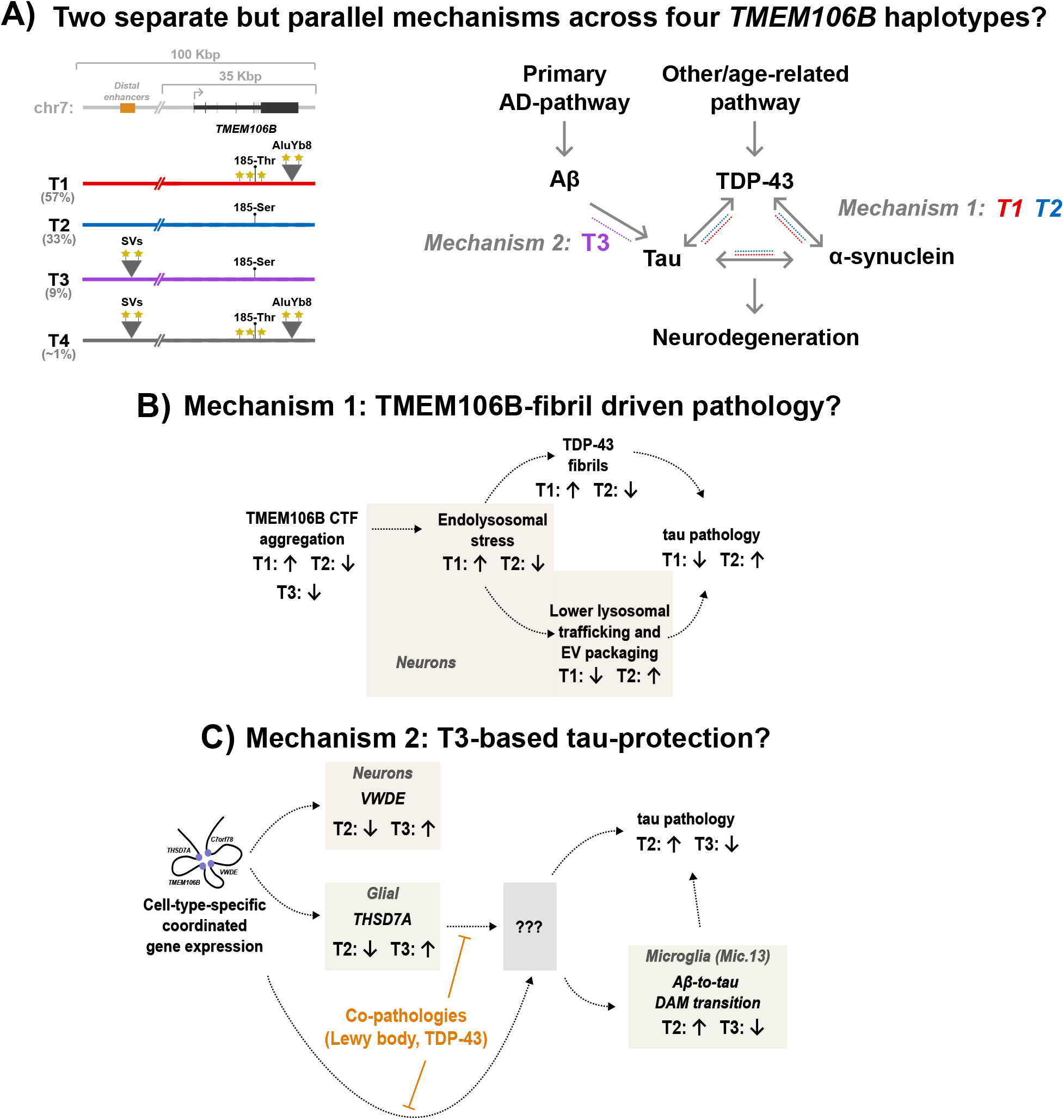
Hypothesized mechanisms underlying the four-haplotype model of *TMEM106B*. **(A)** Schematic overview of the four *TMEM106B* haplotypes and their defining genetic features, alongside Alzheimer’s disease (AD)-related and age-related pathways influencing Aβ, tau, TDP-43 and α-synuclein pathology. We propose that T1 and T2 primarily influence tau, TDP-43 and α-synuclein pathology through an age-related pathway linked to mechanism 1, whereas T3 primarily modulates tau pathology through an AD-related pathway linked to mechanism 2. **(B,C)** Hypothesized mechanisms, functional tests are required. **(B)** Overview of mechanism 1, centered on TMEM106B C-terminal fibril (CTF) abundance. Haplotype effects are shown as upward or downward arrows indicating increased or decreased abundance, respectively; dashed lines indicate no detectable effect. **(C)** Overview of mechanism 2, centered on downstream effects of the T3 haplotype. T2 and T3 show opposing effects on *THSD7A* and *VWDE*, which form a promoter hub with *TMEM106B*. Although the causal drivers of T3-mediated tau assocciation remain unresolved, T3 is associated with reduced prevalence of the Mic.13 microglial state. T3’s association with lower tau pathology is attenuated in the presence of co-pathologies.

In contrast, T3 appears to mark a more favorable resilience axis. Like T2, T3 carries 185-Ser and is associated with lower C-terminal TMEM106B abundance. However, T3 was also associated with reduced tau spread and lower MTBR-tau abundance. This distinguishes T3 from T2, which shares the 185-Ser background but was associated with increased tau pathology, and from T1, whose tau-protective association is accompanied by increased C-terminal TMEM106B abundance and TDP-43 spread. T3 therefore appears to combine TMEM106B-lowering and tau-protective effects without the T1-associated TDP-43 trade-off.

The regulatory mechanisms underlying the T3-specific association with reduced tau pathology remain unclear, as this association was not explained by any single measured feature and may instead reflect one or more upstream regulatory or cellular processes. Potential mechanisms include non-transcriptional effects on TMEM106B protein abundance or broader cis-regulatory effects at the locus, consistent with altered expression of the nearby regulatory-hub genes *THSD7A* and *VWDE*. The association between T3 and reduced Mic.13, a microglial population implicated in AD progression downstream of amyloid and upstream of tau tangle formation, further suggests that T3 may influence the cellular context in which tau pathology emerges. However, because no single transcriptomic, proteomic or microglial feature fully explained the T3-tau association, the protective effect is likely to reflect coordinated effects across multiple regulatory and cellular processes.

In addition to this haplotype-level view, haplotype-tau associations vary by co-pathology context. T3 was associated with reduced tau spread and lower MTBR-tau abundance, and was strongest in settings with limited Lewy body and TDP-43 pathology. This pattern suggests that, as co-pathology accumulates, associations with lower tau pathology may become less strongly linked to T3-associated mechanisms, including partially microglia-related responses, and more strongly linked to neuron-centric endolysosomal pathways associated with T1. However, the T1 association with lower tau pathology was accompanied by increased C-terminal TMEM106B abundance and greater vulnerability to TDP-43 pathology.

Within the studied cohorts, T3 carriage and absence of T1 showed the association profile most consistent with lower combined pathology burden. This model is supported by the depletion of T1 and enrichment of T3 in cognitively healthy centenarians^16^, as well as by genotype-level analyses suggesting that T3 homozygosity may be associated with both tau and TDP-43 pathology. In the context of recent evidence that centenarian resilience involves partial decoupling of amyloid and tau^9^ (Rohde et al. 2026 submitted), T3 may represent one genetic contributor to this decoupling by limiting tau accumulation and spread despite the presence of upstream AD-related pathology.

Several limitations should be considered. Our haplotype-level analysis improves biological resolution but does not identify causal genetic variants or downstream molecular mechanisms. In addition, available proteomic measures identify the soluble fraction of the proteome, but they do not distinguish fibrillar species. Functional studies are required to determine which T3-specific structural or regulatory genomic features underlie its association with lower tau pathology. Furthermore, analyses were performed in cohorts comprising mostly individuals from European-ancestry, and *TMEM106B* haplotypes differ in other ancestries^16^.

Together, our findings identify T3 as a haplotype-level signal associated with lower tau pathology and provide a framework for reconciling the apparently conflicting associations reported by single-variant analyses at the *TMEM106B* locus. These results support a model in which distinct *TMEM106B* haplotypes are associated with partially opposing neurodegenerative traits.

## METHODS AND MATERIALS

### Cohorts and participants

Post-mortem brain characterizations from the 100-plus Study^5^ and the Netherlands Brain Bank^21^, referred to as 100+/NBB (n=193), were previously described^9^. Post-mortem brains from the Religious Order Study and the Memory Aging Project, referred to as ROS/MAP, were previously described^20^. A total of 1,106 ROS/MAP brains were retained after restricting to individuals with: (i) overlapping and quality-controlled genetic data (see below); and (ii) a final cognitive diagnosis of non-demented, mild cognitive impairment, or Alzheimer’s disease dementia (*cogdx* < 6).

All brain donors voluntarily agreed to postmortem brain donation and informed consent was obtained from all brain donors. The protocols of the 100-plus Study protocol and the Netherlands Brain Bank procedures were approved by the Medical Ethics Committee of the Amsterdam UMC. More information on ethics procedures can be found at https://www.brainbank.nl/about-us/ethics/. All participants and/or their legal representatives provided written informed consent for participation in clinical and genetic studies.

### Cohort design and ROS/MAP subset construction

The 100+/NBB collection was retrospectively assembled to maximize contrast in Alzheimer’s disease-related tau pathology while depleting measured age-related co-pathologies, especially TDP-43 and Lewy body^9^. We therefore considered 100+/NBB a design-selected, AD-focused neuropathological context rather than a population-representative aging cohort. To assess whether associations observed in 100+/NBB were also present across broader neuropathological contexts, ROS/MAP was analyzed first in full and subsequently in prespecified nested subsets that progressively approximated the 100+/NBB design.

First, ROS/MAP participants were assigned to 100+/NBB-like strata using NFT-Braak stage and final cognitive diagnosis (*cogdx*): *controls* (Braak ≤ III; no dementia [cogdx *== 1*]), *Alzheimer’s disease cases* (Braak ≥ IV; dementia, [*cogdx ≥ 4*]) and *centenarian* individuals (age at death ≥ 100; no Braak-stage filter; no dementia or mild cognitive impairment [cogdx < 4]). We refer to these three strata as *clinical groups*. Second, within these Braak- and cognition-matched strata, co-pathology-depleted subsets were generated using Lewy body staging (*lewydx_st4*) and TDP-43 staging (*tdp_st4*) to exclude individuals with Lewy body pathology, TDP-43 pathology or both.

The full ROS/MAP analysis therefore estimates haplotype associations in a mixed-pathology aging cohort, whereas the nested analyses assess these associations under increasing alignment with the AD-focused, co-pathology-depleted neuropathological context represented by the 100+/NBB design.

### TMEM106B haplotype genotyping

We used the marker SNPs rs5011439 and rs13237415 to genotype the T1-T4 *TMEM106B* haplotypes in array-imputed data as described in Salazar *et al.*^16^. Array-imputed haplotype data in 100+/NBB were previously described^8^.

For genotyping haplotypes in ROS/MAP, we used Variant Call Format (VCF) data from 1,197 ROS/MAP samples derived from whole-genome sequencing downloaded from Synapse (ID: syn11707419). Data underwent quality control and genotype imputation to guarantee no missing data was present. Briefly, only variants with FILTER=PASS and without multiallelic alleles were retained, and SNPs were restricted to canonical A/C/G/T alleles. Standard variant-level QC was applied, including genotyping rate >99%, minor allele frequency >0.005, or Hardy-Weinberg equilibrium p>1×10^-7^. Individual chromosome data were concatenated; the combined dataset was thinned to 5 million variants (limit for genotype imputation). Genotypes were then lifted over to GRCh38 and prepared for imputation using provided scripts (HRC-1000G-check-bim.pl) specifying TOPMed as reference panel. Finally, all variants were submitted to the TOPMed Imputation server (https://imputation.biodatacatalyst.nhlbi.nih.gov/). The server uses EAGLE (v2.4) (10.1038/ng.3679) to phase data and Minimac4 (10.1038/ng.3656) to perform genotype imputation to the reference panel (version r3). Ancestry analysis was performed by means of clustering with 1000Genome data. Relatedness between individuals was estimated with the kinship coefficient, and unrelated individuals up to third degree were used. Above analyses were performed with PLINK (v2) (10.1086/519795) and custom R scripts. Finally, we used ROS/MAP’s internal sequencing quality control (Synapse ID: syn12178037) to retain samples that pass quality control (QC=Pass) and sex-match (Gender_match=TRUE).

### LC-MS/MS proteomic data

Mass spectrometry-based proteomic analysis for ROS/MAP were based on the final batch-corrected and normalized overall and peptide abundances from Johnson *et al.*^23^, downloaded from Synapse (IDs: syn25006802 and syn52844859, respectively). Abundance for the microtubule binding region of tau (MTBR-tau) was previously estimated^23^ (Synapse ID: syn25006658).

Mass spectrometry-based proteomic analysis of 187 middle temporal gyrus tissues from 100+/NBB (version 2) was performed as previously reported^9^, using the Bruker TIMS-TOF-pro-HT mass spectrometer based soluble fraction of laser-captured MTG grey matter **(Supplementary Methods)**. Normalized peptide-level abundances were estimated using DIA-NN (v1.8.10)^32^ and MS-DAP (v1.2)^33^. These were then used to derive overall protein abundance of THSD7A, TDP-43, and α-synuclein; as well as peptide-specific abundance of the Aβ-portion of the APP protein (**Supplementary Table 3**). For MTBR-tau abundance, as well as overall, N-terminal-, and C-terminal-specific abundance of TMEM106B, we applied Tukey’s median polish^33^ using peptide sequences overlapping each region/domain (**Supplementary Table 3**). We estimated overall cell-type composition across each tissue based on the protein abundance of cell-type markers previously detailed^9^.

We acknowledge that LC-MS/MS measures reflect relative peptide abundance within the analyzed soluble proteomic fraction and do not distinguish full-length proteins from proteolytic fragments, soluble species from fibrillar aggregates, or directly measure fibril burden or seeding activity. Nevertheless, variation in these measures may partly reflect underlying differences in fibrillar protein burden.

### General modeling framework

Motivated by our previous identification of *TMEM106B* haplotypes defined by distinct combinations of coding, structural, and regulatory variation **(Fig. 1b)**^16^, we addressed two overarching research questions: (1) whether T3 shows a distinct association profile with tau and TDP-43 pathology relative to T2, despite both encoding serine at residue 185 (185-Ser); and (2) whether T3 and T1 show concordant or contrasting association patterns across neuropathological and molecular outcomes.

We used regression models to evaluate associations between TMEM106B haplotype dosage and neuropathological or molecular outcomes. Haplotype dosage was defined as the number of copies carried by each individual and was modeled additively. We used three complementary haplotype encodings. First, we used a T2-referenced joint model, where T1, T3, and T4 dosages were included simultaneously, with T2 omitted as the reference haplotype (encodes a Serine at residue 185 and lacks other genetic variants such as large structural variants upstream of the *TMEM106B* gene)^16^. Second, we re-expressed the above joint model as a weighted sum-to-zero encoding, where haplotype coefficients were expressed as deviations from the mean across the modeled haplotypes weighted by their frequency, enabling a direct comparison of their coefficients without selecting a single reference haplotype, described previously^16^. Third, we used a marginal model where each haplotype dosage was evaluated in a separate regression model.

Reflecting the primary research questions, T2-referenced joint model was used for neuropathological and pathology-related outcomes, including NFT-Braak stage, TDP-43 stage, NFT density, MTBR-tau and TDP-43 protein abundance, and microglial-state outcomes. Sum-to-zero joint models were used for both the bulk protein-abundance analyses and single-nucleus transcriptomic analyses of the promoter-hub genes. Marginal models were used for single-nucleus transcriptomic of the endolysosomal gene set. The outcome distribution, analytic datasets, and any additional model-specific covariates are detailed in corresponding Methods sub-sections below.

A common *base covariate set* was applied across analyses and included age at death, sex, amyloid burden, and the first ten genetic ancestry principal components. Additional outcome- and dataset-specific covariates are specified in the corresponding Methods subsections, and include post-mortem interval (PMI), tissue-sample pH, clinical group, study cohort, estimated cell-type composition, local protein abundances, other technical factors.

Amyloid burden was represented using the most appropriate available measure for each cohort and outcome. For NFT-Braak stage in 100+/NBB, we used the amyloid component of the NIA-AA ABC score as a measure of global amyloid burden^34^. In ROS/MAP, immunohistochemistry-derived global amyloid burden (amylsqrt_est_8reg) was used for tau/TDP-43 spread, whereas local amyloid burden (sqt.amyloid_mz) was used for NFT-density, the single-nucleus transcriptomic analyses, and microglial-state analyses; these ROS/MAP models also included PMI. For proteomic analyses in both 100+/NBB and ROS/MAP, local amyloid burden was represented by the abundance of aggregation-prone APP peptides, as described previously^9,23^.

In the 100+/NBB proteomic dataset, tissue pH was missing for five samples. These missing values were imputed using the *mice* package (v3.19.0) in R (v4.5.3), based on available neuropathological measures including amyloid burden, atrophy, atherosclerosis, Lewy body pathology, CAA Thal stage and TDP-43 amygdala positivity. We generated 25 imputed datasets and fitted the proteomic regression models separately in each dataset. Model coefficients and their variances were then pooled across imputations using Rubin’s rules^35^, incorporating both within- and between-imputation variance. Confidence intervals and p-values were calculated from the pooled coefficient estimates and variances rather than by averaging results across imputations.

Unless otherwise specified, statistical significance was defined as a multiple-testing-adjusted p-value < 0.10. This threshold was used for targeted analyses addressing the a priori research questions described above, which were motivated by previously reported *TMEM106B* associations **(Table 1)**. The corresponding Methods subsections specify the correction strategy used for each analysis.

### Associations with tau spread (NFT Braak stage)

To evaluate haplotype associations with tau spread, we organized three complementary analyses frameworks in 100+/NBB and ROS/MAP for analyzing NFT-Braak stage using R (v4.5.3).

*Framework 1 evaluated the overall NFT-Braak-stage distribution*. Braak stage was treated as an ordinal outcome and analyzed using proportional-odds regression implemented with the *clm* function in the ordinal package (v2025.12.29). The model in the complete 100+/NBB collection constituted the primary analysis, and the corresponding model in full ROS/MAP constituted the independent cohort replication. The model was additionally applied to nested ROS/MAP subsets that each progressively approximated the 100+/NBB retrospective selection design.

*Framework 2 localized associations across two spatiotemporally defined transitions in tau spread*. Braak stage was grouped into restricted (0-II), limbic (III-IV), and neocortical (V-VI) involvement, and analyzed using a partial proportional-odds model implemented with *clm*. T1 and T3 were included as nominal terms, allowing their coefficients to vary between restricted versus limbic-or-greater involvement (Braak 0-II versus III-VI) and pre-neocortical versus neocortical involvement (Braak 0-IV versus V-VI). T4 and the model covariates were included as proportional terms. T2 was used as the reference haplotype.

*Framework 3 assessed the robustness of the transition-specific estimates to sparse-data bias*. The same two cumulative transitions from Framework 2 were analyzed using bias-reduced Firth logistic regression implemented in the *logistf* package (v1.26.1). These models were treated as sensitivity analyses of Framework 2.

Consistent with the primary research questions, Benjamini-Hochberg FDR correction was applied separately within each analysis framework and haplotype. In Framework 1, p-values for T3 were pooled across the full 100+/NBB cohort, the full ROS/MAP cohort, and the three nested ROS/MAP subsets and corrected jointly; the same procedure was applied separately to T1. Framework 2 used the same correction strategy, but the number of p-values included in each haplotype-specific corrections doubled because the due to the two spatiotemporal transitions in NFT Braak stage. Framework 3 followed the same correction strategy as Framework 2. T4 was included in all models; in Framework 2, it was constrained to have a proportional association. However, because of its low frequency, T4 was excluded from downstream interpretation and from the multiple-testing correction. Reported q-values therefore control the FDR within each framework- and haplotype-specific family of tests.

### Associations with MTBR-tau abundance

For both 100+/NBB and ROS/MAP, we analyzed overall abundance of the microtubule-binding region of tau (MTBR-tau) derived from LC-MS/MS proteomics using linear regression implemented with *glm* in R. MTBR-tau abundance was z-standardized within each complete proteomic dataset before generating the ROS/MAP subsets. We jointly modeled T1, T3 and T4 haplotype dosage as predictors of MTBR-tau abundance, with T2 serving as the reference haplotype; T4 was retained in the model but excluded from interpretation because of its low frequency. Analyses were performed in 100+/NBB, full ROS/MAP, and the Braak-matched ROS/MAP subset depleted of both Lewy body and TDP-43 pathology (BM, LB-/TDP43-). We selected the latter because it most closely approximated the co-pathology-depleted selection-context of 100+/NBB.

All models included the previously defined base covariate set with additional adjustment for post-mortem interval and clinical group. In 100+/NBB, models further included injection run order, tissue pH and cell-type composition to account for technical and biological factors influencing protein abundance. These covariates were either unavailable in ROS/MAP or already accounted for in the overall and peptide-specific abundance estimates^23^; ROS/MAP models therefore additionally included study cohort (ROS or MAP). Because local TDP-43 and TMEM106B abundance may contribute to variation in tau-related proteomic measures^12–14,31^, we additionally adjusted for the overall local abundance of both proteins to assess whether haplotype associations with MTBR-tau persisted after accounting for these molecular features.

Benjamini-Hochberg FDR correction followed the same haplotype-specific strategy described in the subsection above, with p-values for the T3 coefficients pooled across the 100+/NBB cohort, the full ROS/MAP cohort, and the ROS/MAP BM and LB-/TDP43-subsets and corrected jointly; the same procedure was applied separately to the pooled T1 coefficients.

### Associations with TDP-43 spread and proteomic abundance

TDP-43 spread was evaluated only in ROS/MAP because 100+/NBB was retrospectively selected to minimize co-pathologies, including TDP-43^9^. The ordered TDP-43 staging outcome variable (‘tdp_st4’) was analyzed using the proportional-odds framework described for NFT Braak stage in full ROS/MAP and its Braak-matched, Lewy-body-negative subset.

Z-standardized overall TDP-43 proteomic abundance was analyzed using the linear-regression framework described for MTBR-tau, except that TDP-43 abundance was omitted from the covariate set. We retained TMEM106B abundance and all other applicable dataset-specific covariates. Benjamini-Hochberg correction was similarly applied separately to T3 and T1, with the corresponding p-values pooled across the three datasets and corrected jointly within each haplotype.

### Haplotype associations with microglia states

Prior single-nucleus transcriptomic analyses in ROS/MAP implicated Mic.12 and Mic.13 microglia populations in Alzheimer’s disease progression^27^. Mic.12 has been positioned at an earlier disease stage and linked to amyloid deposition, whereas Mic.13 has been positioned downstream of amyloid and linked more closely to tau accumulation. We therefore first asked whether the T3 association with lower tau pathology was accompanied by lower Mic.13 detection, with or without altered Mic.12 proportional abundance; and then assessed whether T1 showed a concordant or contrasting microglial association pattern.

Because Mic.12 and Mic.13 proportional abundances showed distinct empirical distributions in full ROS/MAP (n=420 individuals with complete genetic and covariate data), models were selected separately for each population. No Mic.13 cells were detected in more than 20% of individuals; Mic.13 detection was therefore analyzed as the primary outcome using logistic regression. Sensitivity analyses examined Mic.13 proportional abundance using fractional logistic regression across all individuals and linear regression restricted to individuals with detected Mic.13 cells **(Supplementary Results)**. Mic.12 was detected in most individuals, and its proportional abundance was analyzed using fractional logistic regression. Haplotype dosages were modeled jointly, with T2 as the reference; T4 was retained as an adjustment term but excluded from interpretation and multiple-testing correction. All models additionally adjusted for study cohort and clinical group. Benjamini-Hochberg correction was similarly applied separately to T3 and T1, with p-values pooled across the two primary microglial outcomes and across the full ROS/MAP cohort and its Braak-matched LB-/TDP43-subset, and corrected jointly within each haplotype.

### Partial correlations of proteomic abundances

We performed a partial correlation analysis of the overall proteomic abundances of Aβ, MTBR-tau, α-synuclein, TDP-43, and TMEM106B. To do this, we used linear regression (*glm* function in R) to regress out the base covariate set along with the defined extended covariates used in the proteomic analyses above. Pairwise Pearson correlations were then calculated between the resulting residuals. Correlations were estimated separately in 100+/NBB and ROS/MAP within three analysis samples: the complete proteomic dataset, the AD stratum comprising AD cases and controls, and the age stratum comprising controls and centenarians. The five proteomic measures yielded ten unique protein pairs within each dataset and subset. Because this analysis was exploratory, all p-values from the ten protein pairs, two complete datasets, and four additional subsets were pooled together and corrected jointly using the Benjamini-Hochberg procedure. Statistical significance was defined as q<0.1.

### Haplotype-based eQTL analysis using single-nucleus RNA-sequencing

We used ROS/MAP single-nucleus transcriptomes from Green *et al.*^27^ to associate the *TMEM106B* haplotypes with two separate gene-sets: (i) *TMEM106B*, *THSD7A*, *VWDE*, and *C7orf78*, which possibly forms a four-gene promoter hub in neurons based on publicly available PLAC-seq data **(Fig. 4b)**^24^; and (ii) genes known to be involved in endolysosomal-related pathways (see below).

First, we used the cell-atlas from Green *et al.* (n=437, Synapse ID: syn53694215) and applied the *d-sum* normalization procedure from Cumo *et al.* 2021^36^ to each of the cell-type-specific Seurat objects downloaded from Synapse (ID: syn53366818). Briefly, the d-sum procedure summarises single-nucleus data at the donor level for each cell type within each Seurat (v5.3.0) object^37^, and we retained only cell types represented by at least 10 donors, each contributing ≥10 cells. For each cell type, raw counts were summed per donor using Seurat’s *AggregateExpression* function, producing donor-level count matrices that were formatted in a *DGEList* from the edgeR package (v4.6.2). Genes were automatically filtered for sufficient expression using edgeR’s *filterByExpr* function, described in detail by Chen *et al.*^38^. The retained genes were normalized by edgeR’s *TMM* (Trimmed Mean of M-values) function to correct for library size differences, then transformed to log2-counts per million (log2-CPM) using the *voom* function from the limma package (v3.64.1), followed by a rank-based inverse normal transformation across donors for each gene. We retained samples that overlap with QC-filtered genetics data (see above), resulting in a total of 420 samples.

To associate *TMEM106B* haplotypes with expression of genes in the four-gene promoter hub, we first fit a joint regression model using the *glm* function in R, including all modeled haplotypes simultaneously with T2 as the reference haplotype. Haplotype effects were subsequently re-expressed using sum-to-zero encoding (see subsection above). Models were fit separately for each available promoter-hub gene, and adjusted for the base covariate set along with additional possible single-nucleus-specific confounders: clinical group, study cohort, and sequencing library-related variables (‘Total_Genes_Detected’ and ‘Estimated_Number_of_Cells’).

These targeted secondary analyses were designed to characterize and compare molecular association patterns across the three common *TMEM106B* haplotypes. Accordingly, all p-values from the joint associations of all haplotypes with the detectably expressed promoter-hub genes across seven cell types were pooled and corrected together. Because these tests were linked due to overlapping haplotypes, promoter-hubs, and shared individuals across cell-types, we estimated the effective number of independent tests (*Meff*) as described by Li and Ji^25^. We then used the estimated *Meff* value to adjust for multiple testing using Bonferroni correction, with statistical significance defined as an adjusted p-value < 0.1. T4 was included in the models but excluded because of its low frequency.

To explore broader transcriptional associations of the haplotypes within the endolysosomal system, we curated an endolysosomal-based gene-set using MSigDB^39,40^. To do this, we used the *msigdbr* package (v25.1.1) in R to query genes involved in lysosome, endosome, endocytosis, phagocytosis, and autophagy by using the regex string, “LYSO|ENDOSOM|ENDOCY|PHAGOCY|PHAGY”, to search in the following collections: pathways (category “H”), molecular function (category “C5”), biological process (category “C5”), cellular component (category “C5”), and reactome (category “C2”). We then excluded all possible ribosomal or mitochondrial genes based on the following regex string, “RIBOSOM|TRANSLATION|MITOCHON”, queried on the same collections, and retained those that were expressed in at least 66% of all samples (n=277). This generated a unique gene-set of 1,383 genes **(Supplementary Table 1)**.

We then used marginal regression models to evaluate associations between *TMEM106B* haplotypes and cell type-specific expression of genes in the endolysosomal gene set, using the *glm* function in R and adjusting for the same covariates as in the promoter-hub transcriptomic analyses. Given the exploratory and high-dimensional nature of this analysis, all p-values were pooled across haplotypes, genes, and cell-types and corrected jointly using the Benjamini-Hochberg procedure, with statistical significance defined as q<0.01. T4 was excluded because of its low frequency.

### Associations with overall and domain-specific abundance of TMEM106B and THSD7A

To assess whether the promoter-hub transcriptomic associations extended to protein abundance, and whether T3 and T1 showed concordant or contrasting protein-level profiles, we analyzed overall TMEM106B and THSD7A abundance, and N- and C-terminal TMEM106B peptide abundance in 100+/NBB, full ROS/MAP, and the ROS/MAP BM, LB-/TDP43-subset. For each abundance, we used the weighted sum-to-zero encoding model to compare associations between the haplotypes (see subsection above). Applicable base and dataset-specific proteomic covariates were included as described in the subsections above. Because these targeted protein analyses aimed to compare molecular association profiles of the haplotypes, we pooled all p-values from all haplotypes in the overall protein abundance analyses (TMEM106B and THSD7A) and corrected jointly using Benjamini-Hochberg procedure. The same approach was done for correcting p-values in the domain-specific abundance analyses (N-terminal and C-terminal). Statistical significance was defined as q<0.10. T4 was included in all models, but excluded from interpretation and multiple-testing correction because of its low frequency.

## DATA AVAILABILITY

ROS/MAP clinical and neuropathological data are available through the Rush Alzheimer’s Disease Center Research Resource Sharing Hub (https://www.radc.rush.edu/) under an approved data-use agreement. Molecular and omics data are available through Synapse under accession syn3219045 (https://www.synapse.org/Synapse:syn3219045), subject to the applicable access requirements. Data from the 100-plus Study are available through the Alzheimer Genetics Hub (https://alzheimergenetics.org/) upon reasonable request. Netherlands Brain Bank data are available through the Netherlands Neurogenomics Database (https://nnd.academy/) or upon reasonable request.

## ACKNOWLEDGEMENTS

We thank all participating centenarians and their family members, as well as all other brain donors. We acknowledge the staff of the Netherlands Brain Bank and the Rush Alzheimer’s Disease Center Research Resource Sharing Hub for their cooperation. We also acknowledge all current and former members of the 100-plus Study practical team who contributed to data collection. We thank Gilad Green for valuable input on the ROS/MAP single-nucleus transcriptomic data and Anke A. Dijkstra for valuable input on neuropathological interpretation.

## AUTHOR CONTRIBUTIONS

A.N.S. and H.H. conceived and designed the study. F.K., K.W.L. and A.B.S. generated and curated the 100+/NBB proteomic data, with M.H. contributing proteomic expertise. S.K.R. and M.C.L. generated and curated the 100+/NBB neuropathological data, with A.J.M.R. contributing neuropathological expertise. A.N.S. and N.T. curated the genetic data, with S.vd.L. contributing genetic expertise. A.N.S. performed the statistical and computational analyses and interpreted the results. A.N.S. and H.H. wrote the manuscript. H.H. and S.vd.L. supervised the study and acquired funding.

