## Supplementary Information for "*TMEM106B* haplotypes show distinct associations with tau and TDP-43 pathologies in the aging brain"

### SUPPLEMENTAL RESULTS

#### *Sensitivity analysis of the haplotype associations with Mic.12 and Mic.13 microglia states*

To evaluate the robustness of the Mic.13 association to model specification, we performed additional sensitivity analyses using fractional logit regression of overall proportional abundance and Gaussian linear models restricted to individuals with detectable Mic.13 cells. Fractional logit models demonstrated directionally concordant associations for both T1 and T3, with T3 retaining its effect size with reduced Mic.13 abundance ($\beta=-0.26$, 95% CI= [−0.59,0.06], $p=0.124$). T1 showed a similar trend in the same direction ($\beta=-0.19$, 95% CI=[−0.39,0.00], $p=0.050$), whereas the corresponding logistic regression association was not statistically significant (OR=0.78, 95% CI =[0.51,1.18], $p=0.24$). In contrast, among individuals with detectable Mic.13 cells, proportional abundance did not differ by either T3 genotype ($\beta=-0.006$, 95% CI =[−0.020,0.008],$p=0.401$) or T1 genotype ($\beta=-0.006$, 95% CI=[−0.014,0.002], $p=0.117$). Together, these analyses suggest that the association with Mic.13 primarily reflects differences in the probability of detecting the cell state, rather than differences in its proportional abundance among individuals in whom Mic.13 is detected.

### SUPPLEMENTAL METHODS

#### *Mass spectrometry proteomic analysis*

DIA-PASEF raw data were processed with DIA-NN 1.8.1. An in-silico spectral library was generated from the uniprot human proteome (SwissProt and TrEMBL, canonical and additional isoforms, release 2024-02) using Trypsin/P digestion and at most 1 missed cleavage. Fixed modification was set to carbamidomethylation(C) and variable modifications were oxidation(M) and N-term M excision (at most 1 per peptide). Peptide length was set to 7-30, precursor charge range was set to 2-4, precursor m/z was limited to 280-1220, both MS1 and MS2 mass accuracy were set to 15 ppm, double-pass-mode and match-between-runs were enabled. Protein identifiers (isoforms) were used for protein inference. All other settings were left as default.

MS-DAP 1.2 was used for downstream analyses of the DIA-NN results. Peptide-level filtering was configured to retain only peptides that were confidently identified in at least 50% of samples in at least 1 of the sample groups (NDC, ADC, CHC). Only proteins with at least 2 peptides were retained. Peptide abundance values were normalized using the MWMB algorithm, followed by protein-level mode-between normalization. 9 outlier samples were identified in the quality control analyses presented in the MS-DAP report, which we subsequently excluded from statistical analyses.

SUPPLEMENTAL FIGURES

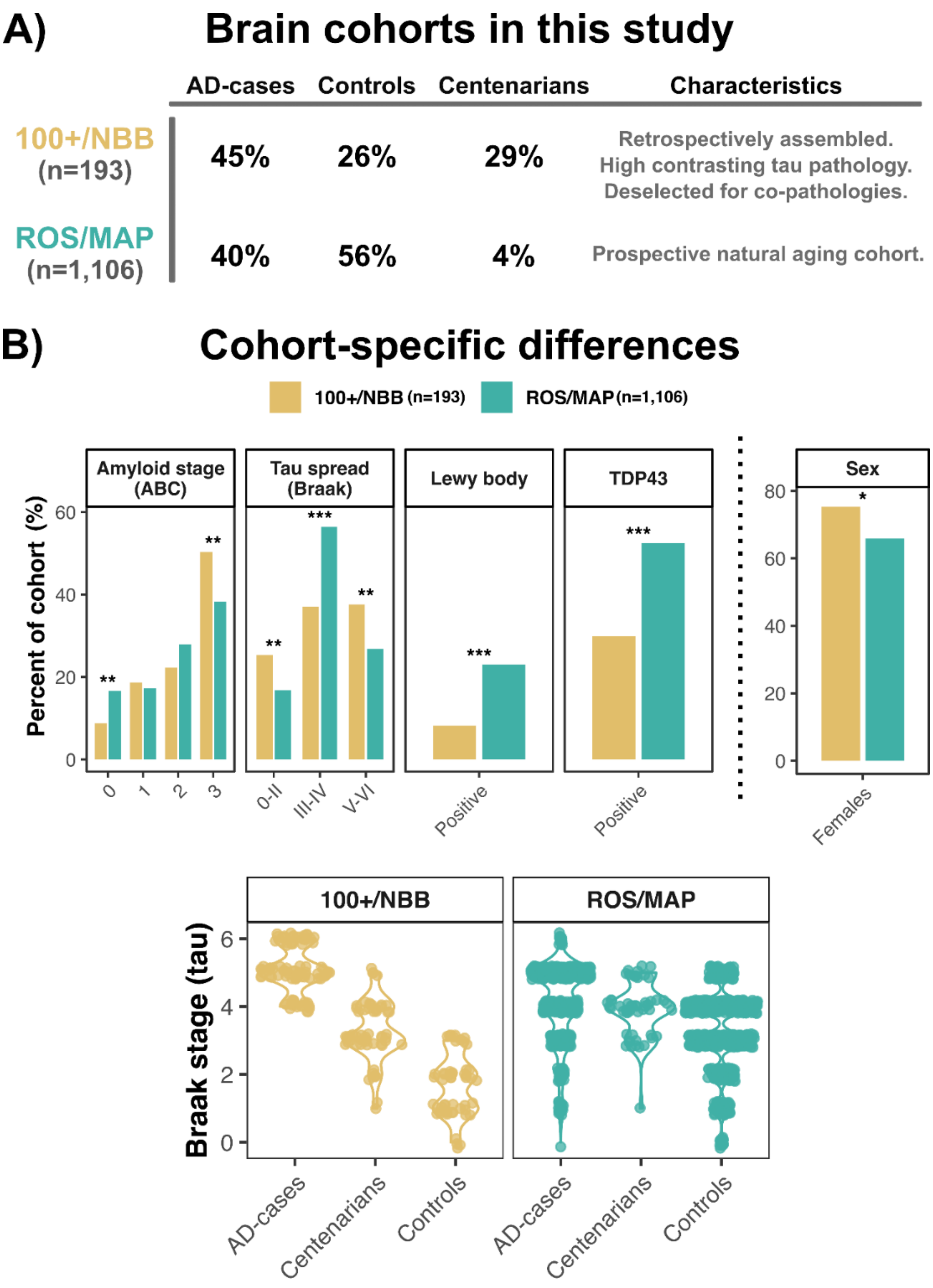


**Supplementary Fig. 1 | Neuropathological and epidemiological features of the 100+/NBB and ROS/MAP brain-sample collections. (A)** Distribution of AD cases, controls, and centenarians. **(B)** Top: neuropathology and sex distributions. FDR-adjusted p (5% threshold): *** < 0.001; ** < 0.01; * < 0.05. Bottom: Braak stage distributions.

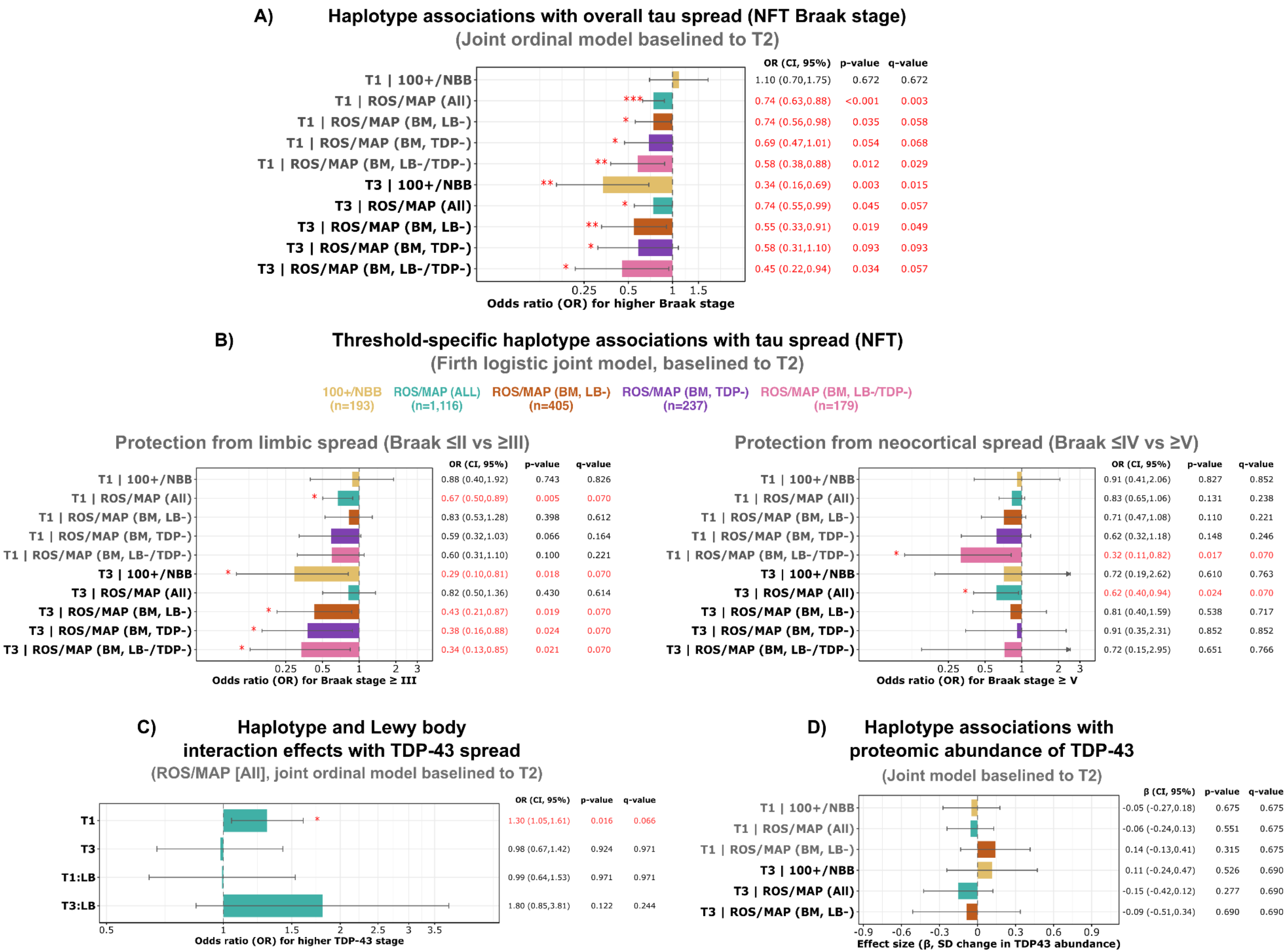
A) Haplotype associations with overall tau spread (NFT Braak stage)
(Joint ordinal model baselined to T2)
OR (CI, 95%) p-value q-value
T1 | 100+/NBB 1.10 (0.70,1.75) 0.672 0.672
T1 | ROS/MAP (All) *** 0.74 (0.63,0.88) <0.001 0.003
T1 | ROS/MAP (BM, LB-) * 0.74 (0.56,0.98) 0.035 0.058
T1 | ROS/MAP (BM, TDP-) * 0.69 (0.47,1.01) 0.054 0.068
T1 | ROS/MAP (BM, LB-/TDP-) ** 0.58 (0.38,0.88) 0.012 0.029
T3 | 100+/NBB ** 0.34 (0.16,0.69) 0.003 0.015
T3 | ROS/MAP (All) * 0.74 (0.55,0.99) 0.045 0.057
T3 | ROS/MAP (BM, LB-) ** 0.55 (0.33,0.91) 0.019 0.049
T3 | ROS/MAP (BM, TDP-) * 0.58 (0.31,1.10) 0.093 0.093
T3 | ROS/MAP (BM, LB-/TDP-) * 0.45 (0.22,0.94) 0.034 0.057
0.25 0.5 1 1.5
Odds ratio (OR) for higher Braak stage
B) Threshold-specific haplotype associations with tau spread (NFT)
(Firth logistic joint model, baselined to T2)
100+/NBB (n=193) ROS/MAP (ALL) (n=1,116) ROS/MAP (BM, LB-) (n=405) ROS/MAP (BM, TDP-) (n=237) ROS/MAP (BM, LB-/TDP-) (n=179)
Protection from limbic spread (Braak ≤II vs ≥III)
OR (CI, 95%) p-value q-value
T1 | 100+/NBB 0.88 (0.40,1.92) 0.743 0.826
T1 | ROS/MAP (All) * 0.67 (0.50,0.89) 0.005 0.070
T1 | ROS/MAP (BM, LB-) 0.83 (0.53,1.28) 0.398 0.612
T1 | ROS/MAP (BM, TDP-) 0.59 (0.32,1.03) 0.066 0.164
T1 | ROS/MAP (BM, LB-/TDP-) 0.60 (0.31,1.10) 0.100 0.221
T3 | 100+/NBB * 0.29 (0.10,0.81) 0.018 0.070
T3 | ROS/MAP (All) 0.82 (0.50,1.36) 0.430 0.614
T3 | ROS/MAP (BM, LB-) * 0.43 (0.21,0.87) 0.019 0.070
T3 | ROS/MAP (BM, TDP-) * 0.38 (0.16,0.88) 0.024 0.070
T3 | ROS/MAP (BM, LB-/TDP-) * 0.34 (0.13,0.85) 0.021 0.070
0.25 0.5 1 2 3
Odds ratio (OR) for Braak stage ≥ III
Protection from neocortical spread (Braak ≤IV vs ≥V)
OR (CI, 95%) p-value q-value
T1 | 100+/NBB 0.91 (0.41,2.06) 0.827 0.852
T1 | ROS/MAP (All) 0.83 (0.65,1.06) 0.131 0.238
T1 | ROS/MAP (BM, LB-) 0.71 (0.47,1.08) 0.110 0.221
T1 | ROS/MAP (BM, TDP-) 0.62 (0.32,1.18) 0.148 0.246
T1 | ROS/MAP (BM, LB-/TDP-) * 0.32 (0.11,0.82) 0.017 0.070
T3 | 100+/NBB 0.72 (0.19,2.62) 0.610 0.763
T3 | ROS/MAP (All) * 0.62 (0.40,0.94) 0.024 0.070
T3 | ROS/MAP (BM, LB-) 0.81 (0.40,1.59) 0.538 0.717
T3 | ROS/MAP (BM, TDP-) 0.91 (0.35,2.31) 0.852 0.852
T3 | ROS/MAP (BM, LB-/TDP-) 0.72 (0.15,2.95) 0.651 0.766
0.25 0.5 1 2 3
Odds ratio (OR) for Braak stage ≥ V
C) Haplotype and Lewy body interaction effects with TDP-43 spread
(ROS/MAP [All], joint ordinal model baselined to T2)
OR (CI, 95%) p-value q-value
T1 * 1.30 (1.05,1.61) 0.016 0.066
T3 0.98 (0.67,1.42) 0.924 0.971
T1:LB 0.99 (0.64,1.53) 0.971 0.971
T3:LB 1.80 (0.85,3.81) 0.122 0.244
0.5 1 1.5 2 2.5 3 3.5
Odds ratio (OR) for higher TDP-43 stage
D) Haplotype associations with proteomic abundance of TDP-43
(Joint model baselined to T2)
β (CI, 95%) p-value q-value
T1 | 100+/NBB -0.05 (-0.27,0.18) 0.675 0.675
T1 | ROS/MAP (All) -0.06 (-0.24,0.13) 0.551 0.675
T1 | ROS/MAP (BM, LB-) 0.14 (-0.13,0.41) 0.315 0.675
T3 | 100+/NBB 0.11 (-0.24,0.47) 0.526 0.690
T3 | ROS/MAP (All) -0.15 (-0.42,0.12) 0.277 0.690
T3 | ROS/MAP (BM, LB-) -0.09 (-0.51,0.34) 0.690 0.690
-0.9 -0.6 -0.3 0 0.3 0.6 0.9
Effect size (β, SD change in TDP43 abundance)

**Supplementary Fig. 2 | Extended haplotype associations with tau and TDP-43 spread and abundance. (A-E)** Associations between *TMEM106B* haplotype carriership and tau or TDP-43 across all post-mortem brain collections and subsets, each with a unique color as defined in Fig. 1C. Colored bars indicate estimated odds ratio (OR) or beta coefficient, and black horizontal lines represent 95% confidence intervals. Asterisks indicate FDR-adjusted q-values (10% threshold): *** $q < 0.01$; ** $q < 0.05$; * $q < 0.1$. Red text denotes an association with statistical significance ($q<0.1$) **(A)** Joint ordinal model with overall NFT Braak stage. **(B)** Joint biased-reduced Firth logistic regression model testing threshold-specific haplotype effects on tau spread: protection from early limbic spread (left; Braak ≤II vs ≥III) and protection from neocortical spread (right; Braak ≤IV vs ≥V). **(C)** Interaction effects in a joint ordinal model with overall TDP-43 spread. **(D)** Joint linear model on TDP-43 protein abundance based on proteomic datasets (see Fig. 3A).

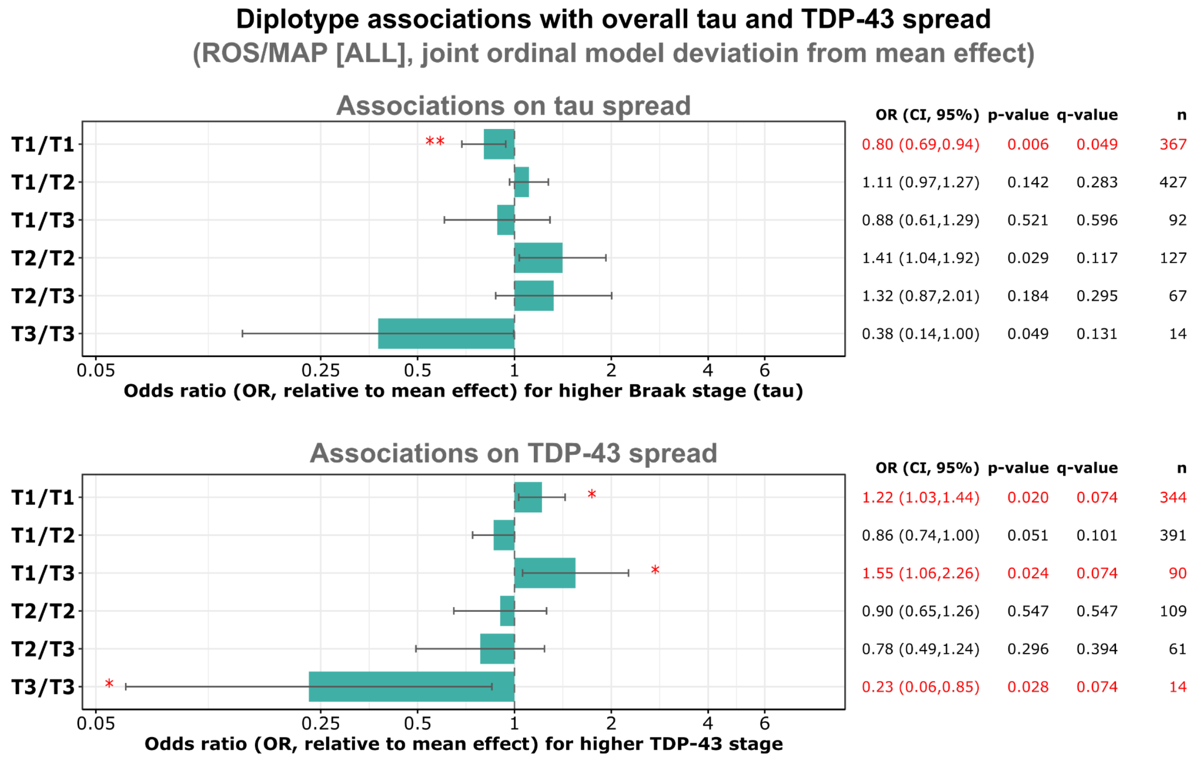


**Supplementary Fig. 3 | Haplotype combination (diplotype) associations with tau and TDP-43 stage in ROS/MAP.** Effects are shown as odds ratios derived from sum-to-zero encoded models, representing deviations relative to the mean effect across haplotypes. Top: effect on tau spread (NFT Braak stage). Bottom: effect on TDP-43 spread (TDP-43 stage). Asterisks represent FDR-adjusted q-value (10% threshold): *** < 0.01; ** < 0.05; * < 0.1. Red indicates statistical significance (q-value < 0.1).
